# Assessment of antibody responses to *Anopheles* SG6-P1 and *Aedes* N-term 34kDa salivary peptides: a human-challenge trial of controlled exposures to vector bites

**DOI:** 10.1101/2025.06.16.25329674

**Authors:** Victor Chaumeau, Ellen A. Kearney, Praphan Wasisakun, Sunisa Sawasdichai, Aye Aye Aung, Paul A. Agius, Thaw Htwe Min, Daniela da Silva Gonçalves, Katherine O’Flaherty, Angeline Rouers, Sarang Aryalamloed, Gay Nay Htoo, Mu Phang Sue, Naw Moo Tha, Naw Chanida, Gornpan Gornsawun, Sadudee Chotirat, Julie A. Simpson, Laurent Rénia, François Nosten, Freya J. I. Fowkes

## Abstract

**Background:** Human antibodies against mosquito salivary proteins are proposed as proxy biomarkers of exposure to vector bites. This trial sought to characterise the boosting and decay dynamics of antibodies against *Anopheles* SG6-P1 and *Aedes* N-term 34kDa salivary peptides in a human challenge model of controlled exposure to the main Southeast Asian malaria and global dengue vectors.

**Methods:** In this single-centre, open-label, randomised, exploratory factorial trial, healthy volunteers aged 18-60 years with no history of recent travel to rural areas were recruited in Mae Sot, Thailand (ClincalTrials.gov: NCT04478370). Participants were randomly assigned to receive either 35 or 305 bites of mosquitos of laboratory-adapted colonies of *An. dirus*, *An. maculatus*, *An. minimus*, *Ae. aegypti*, and *Ae. albopictus* using a block randomisation schedule. Samples were collected weekly before, during and after the challenges for 16 weeks. The primary endpoint was total IgG antibodies against *Anopheles* SG6-P1 peptides measured using high-throughput ELISA and analysed with Generalized Estimating Equations. Outcome assessors were masked to the intervention groups.

**Findings:** Between January 21, 2021, and May 10, 2022, 248 volunteers were screened, of whom 210 were randomly assigned to receive either 35 or 305 bites of *Ae. aegypti* (n=20 and n=19, respectively), *Ae. albopictus* (n=20, n=21), *An. dirus* (n=21, n=21), *An. maculatus* (n=23, n=24), or *An. minimus* (n=22, n=19), comprising the intention-to-treat population. In participants exposed to 305 *An. minimus* bites, total anti-gSG6-P1 IgG levels increased 1.14-fold (95%CI: 1.03-1.26) and 1.18-fold (95%CI: 1.05-1.33) during the exposure and post-exposure periods respectively (relative to baseline), with minimal or no boosting observed in other groups. The estimated half-life of anti-gSG6-P1 antibodies was 421 (95%CI: 155-688) days. Seven participants were withdrawn due to an adverse event.

**Interpretation:** Anti-gSG6-P1 antibodies were boosted in response to exposure to 305 bites of *An. minimus* but the magnitude of boosting was small and antibodies decayed slowly. Future research is warranted to identify and validate serological markers of vector biting exposures.

**Funding:** Wellcome Trust, NHMRC.

**Research in Context:** *Evidence before this study:* Human antibodies against mosquito salivary proteins have been investigated as serological biomarkers of exposure to bites of mosquitos that transmit malaria (*Anopheles*) and dengue (*Aedes*), however their associations with and dynamics following biting exposures remain unknown. On June 3 2020, we searched published articles in PubMed and MEDLINE using the search terms ((Anophel* OR Aede*) AND saliva* AND (antibod* OR sero* OR antigen OR marker* OR biomarker*)). We systematically reviewed studies investigating the *An. gambiae* salivary gland protein 6 (gSG6), its derivative gSG6-P1 peptide, or the *Ae. aegypti* N-term 34kDa peptide as an outcome measure of biting exposures. We used multilevel modelling to assess the association between population-level anti-gSG6 IgG antibody seroprevalence and *Anopheles* human-biting rates reported in 12 studies. The results showed that seroprevalence and vector biting rates are positively associated and that this association is stronger in African settings where *An. gambiae* is the only dominant vector species than in areas where *An. gambiae* is absent. Five studies investigated anti-*Ae. aegypti* N-term 34kDa IgG antibody responses but the associations with *Aedes* biting rate were not assessed. This review also identified a knowledge gap on the association between antibodies against either gSG6 or *Ae. aegypti* N-term 34kDa and human-biting rates measured at the individual level or under conditions of controlled exposure to accurately quantify boosting and decay dynamics.

*Added value of the study:* This is the first assessment of boosting and decay dynamics of antibody responses directed against mosquito salivary antigens in a human challenge model of controlled exposure to vector bites. Small boosts of long-lasting antibodies directed against gSG6-P1 and *Ae. aegypti* N-term 34kDa peptides, as well as orthologous peptides designed using published sialomes of *An. minimus*, *An. maculatus*, *An. dirus* and *Ae. albopictus*, were detected in response to the study challenges. This innovative trial design allows determination of the dose-response relationship between mosquito biting exposures and antibody responses, the rate of antibody decay, and the cross-reactivity of anti-salivary antibody responses across species of biting exposure, thereby providing crucial information for the validation of antibodies against mosquito saliva as a quantitative outcome measure of recent human-vector contact.

*Implications of the available evidence:* This study demonstrates minimal boosting and slow decay of antibodies to *Anopheles* SG6-P1 and *Aedes* N-term 34kDa salivary antigens following controlled biting exposures in a cohort of participants with relatively high levels of baseline seroreactivity against those peptides. These findings suggest somewhat limited utility of anti-salivary antibodies to measure changes in individual-level biting exposures from these mosquito species over short periods of time, however, their utility to measure population level exposure over longer periods of time is yet to be determined. This will impact how these serological biomarkers can be applied to evaluate the effectiveness of vector control interventions or for serosurveillance, whereby larger samples sizes or longer follow up may be required to accurately capture boosting and decay dynamics.

## Introduction

Mosquitos are major vectors that cause approximately 350 million annual cases of important diseases, including malaria, dengue and chikungunya^1^. While there has been significant progress in reducing the burden of malaria in Southeast Asia in recent decades, increasing heterogeneity in transmission as cases decline is a barrier to achieving elimination. Surveillance of the vector populations has therefore been identified as a priority to monitor complex transmission dynamics and inform receptivity. The main vectors in this region are *Anopheles dirus*, *An. maculatus*, and *An. minimus*; several other species are secondary vectors^2^. These species exhibit early and outdoor biting behaviours that limit the efficacy of mosquito bed nets^3^. Dengue viruses are transmitted by aedine mosquitoes. *Aedes aegypti* and *Ae. albopictus* are the main vectors globally^4^. Infection can cause severe disease and death and the overall burden has increased over the past decades^5^. As there is no specific treatment and no safe, effective, and widely available vaccine, prevention of infection with vector-control and personal protection is critical to reduce morbidity and mortality.

Human exposure to vector bites is an important parameter of vectorial capacity and transmission models^6^. Measuring these exposures is therefore essential to assess the dynamics of disease transmission and evaluate the efficacy of vector control and transmission-blocking interventions. However, there is no tool to measure mosquito biting exposures accurately and at scale^7^. This knowledge gap constitutes an important barrier to disease control and elimination.

Upon blood feeding, female mosquitos inject their saliva in the skin of vertebrates. Mosquito saliva is composed of hundreds of biologically active components that play essential roles in the physiology of blood feeding and elicit immune responses in vertebrates^8^. Assessment of antibodies directed against mosquito saliva in human blood samples is proposed as a surrogate measure of exposures to bites of medically important mosquito vector species^9^. Using omics approaches, dozens of *Anopheles*- and *Aedes*-specific salivary proteins have been discovered thereby opening avenues for the development of biomarkers specific of vector biting exposures^10,11^. Initial investigations identified antibody responses to *An. gambiae* salivary gland protein 6 (gSG6) and its peptide derivative (gSG6-P1) in individuals naturally bitten by *Anopheles* mosquitos in Africa^12,13^. Similar investigations have identified the *Aedes aegypti* N-term 34kDa (aeg34kDa) salivary peptide as a potential serological biomarker of exposure to *Aedes* bites^14^. Subsequently, measurement of human antibody responses to these salivary antigens have been used as a population-level outcome to assess vector biting exposures, estimate risk of disease transmission, and to evaluate vector control interventions globally^14–16^. To date the leading candidate salivary antigens have primarily been investigated as biomarkers of species-specific biting exposure, e.g. gSG6 to measure *An. gambiae* exposure and aeg34kDa to measure *Ae. aegypti* exposure. Indeed, a systematic review with pooled analysis found that when applied in settings outside Africa where *An. gambiae* is absent, the association between human biting rates and anti-gSG6 IgG was weaker^15^. A study on La Reunion Island (where only *Ae. albopictus* is present) identified a positive association with antibodies against aeg34kDa and biting exposure^16^. This highlights a need for further investigations to quantify the effect of biting exposures from other *Anopheles* and *Aedes* species on the anti-gSG6 and anti-aeg34kDa antibody response. Additionally, this suggests a potential role for novel peptide orthologs to serve as species-specific markers of exposure to bites of *Ae. albopictus* and *Anopheles* species that predominate in regions outside of Africa. A species-specific approach may help overcome limitations of varied sequence identity between the SG6 and N-term 34kDa proteins of *An. gambiae* and *Ae. aegypti* with other major *Anopheles* and *Aedes* species, respectively^10,11^. This approach is being trialled to identify novel non-SG6 salivary antigens in South America^18,19^, where the SG6 gene is absent from the dominant vectors (*An. darlingi* and *An. albimanus*), however, it is yet to be trialled in Southeast Asia where the sequence identity to gSG6 ranges from 52-78% across the dominant vectors^10^.

There is a key knowledge gap surrounding individual-level boosting and decay dynamics of anti-salivary antibodies following exposure to mosquito bites which prevents their scale up for these proposed applications. This gap is primarily due to inherent challenges in observing an individual’s total mosquito biting exposure over time in natural settings, which may be better simulated under conditions of controlled exposure. To our knowledge, only one study has sought to characterise the individual-level longitudinal antibody responses following controlled human exposure to ∼100 *Culex quinquefasciatus* bites fortnightly for 10 months^17^. IgG antibodies to *Cx. quinquefasciatus* salivary extracts were measured 4-weekly and were found to boost and decay rapidly before being sustained across the remainder of the study, potentially suggesting the development of a tolerance. However, further studies would be required to confirm these observations as the sample size for this study was only one participant who had no prior exposure to *Cx. quinquefasciatus* mosquito bites. An expanded version of this study design that captures rich data on the number bites received and the antibody response measured at the individual level could be useful to characterise the dynamics and dose-response relationship of antibody response to *Anopheles* and *Aedes* salivary antigens following biting exposures.

The aim of this study was to assess the boosting and decay dynamics of total IgG responses directed against the *Anopheles* SG6-P1 and *Aedes* N-term 34kDa salivary peptides in a human challenge model of controlled exposure to bites of *An. dirus*, *An. maculatus*, *An. minimus* (the main malaria vectors in the Greater Mekong Subregion), *Ae. aegypti*, and *Ae. albopictus* (the main dengue vectors worldwide).

## Methods

### Study design

This study is an exploratory, factorial randomized controlled trial of exposure to mosquito bites with 10 arms corresponding to different species (*An. minimus*, *An. maculatus*, *An. dirus*, *Ae. aegypti* and *Ae. albopictus*) and numbers of bites (35 or 305 bites in total split in 7 weekly challenges over 6 weeks). The study was conducted at the Shoklo Malaria Research Unit in Mae Sot, Thailand. The protocol, participant information sheet and informed consent form were approved by the Oxford Tropical Research Ethics Committee (reference 62-19), the Ethics Committee of the Faculty of Tropical Medicine, Mahidol University (reference TMEC 20-007), the Alfred Hospital Ethics Committee (reference 630/21) and the Tak Community Advisory Board (reference TCAB202003), a community-based committee assembling members of the communities in which the study was performed^20^. The trial was registered in ClinicalTrials.gov (NCT04478370) and the detailed protocol was published separately^21^.

### Participants

Participants were generally healthy individuals aged 18 to 60 years old as assessed by a medical doctor, of Thai, Burmese or Karen ethnicity, living in Mae Sot city for the last 12 months and able to tolerate mosquito biting exposures. Exclusion criteria included individuals with a history of travel in a rural area (i.e., may be exposed to *Anopheles* bites) in the last 12 months, or plan to do so during the study, medication or condition deemed to interfere with the outcome measure or increase the risk of an adverse reaction to the study procedures, and haemoglobin concentration less than 110 g/L of blood. Pregnant and breastfeeding women were also excluded. Information on the study was spread through word of mouth by the study team to people who live in Mae Sot and individuals interested in participating were invited to contact the study team. To reach the planned sample size, interested volunteers were asked to further spread the study information amongst their networks. Gender was self-reported by the participants and every answer provided by them was recorded. All participants gave their written informed consent for participating in the study.

### Randomisation and masking

A block randomization schedule was generated using the variables species (*An. minimus*, *An. maculatus*, *An. dirus*, *Ae. aegypti* and *Ae. albopictus*) and dose (35 or 305 bites in total), yielding an ordered list of 15 blocks with 10 participants per block randomly assigned to one of the 10 study arms. An allocation sequence was implemented using individual, sealed and sequentially numbered envelopes. Following screening and eligibility assessment, participants were assigned to a study arm during visit two using the randomization schedule. The allocation sequence was generated by a study investigator. At the beginning of the study, the study coordinator prepared a set of case report forms with pre-printed subject identification codes and attached the sealed envelope containing intervention allocation to the case report forms. Then, the study nurses assigned a subject identification code to participants by chronological order of enrolment in the study and the envelope was opened during visit two. Allocation of intervention was masked to outcome assessors (laboratory personnel who processed the serum samples). In order to do so, allocation of intervention in study datasets that contain this information was masked until all antibody data were made available to study investigators and statisticians.

### Procedures

Following screening and eligibility assessment, participants were appointed to attend weekly visits for 16 weeks and the complete follow-up was 112 days. Participants were challenged with bites of laboratory-adapted colonies of *An. minimus*, *An. maculatus*, *An. dirus*, *Ae. aegypti* or *Ae. albopictus* weekly between day 14 and day 56 (7 times in total). Participants in the low-exposure arms were challenged on each occasion with five mosquito bites (35 bites in total), those in the high-exposure arms were challenged once with five bites and then six times with 50 bites (305 bites in total). Immediate skin reactions were recorded 20 to 30 minutes after every challenge and delayed skin reactions were recorded 24 to 36 hours after the first and second challenges. The level of antibody titres against mosquito salivary antigens was measured in participant serum from venous blood collected at one-week intervals for the entire follow-up.

### Immunological assays

The peptide panel tested in this study included the gSG6-P1 peptide (derived from the SG6 protein sequence of *An. gambiae* and originally designed by Poinsignon *et al.*^13^), the corresponding orthologous peptides dirSG6-P1, macSG6-P1, and min-SG6-P1 (designed for this study using the SG6 protein sequence of *An. dirus*, *An. maculatus* and *An. minimus* published previously^10^), the N-term 34 kDa peptide of *Ae. aegypti* (originally designed by Elanga Ndille *et al.*^14^) and the corresponding orthologous peptide in *Ae. albopictus* also designed in-house (the details of peptide design and sequences are provided in Appendix). Total IgG antibodies against these peptides were measured by adapting previously published ELISA protocols^13,16,22,23^, and optimising them into a high-throughput platform (detailed in Appendix). Antibody response is reported as levels (measured as optical density [OD]) and seropositivity (defined for each peptide as having an OD greater than the mean plus three standard deviations of the unexposed Melbourne controls).

### Outcomes

The primary outcome of this study was the total IgG antibody responses to the *Anopheles* SG6-P1 salivary peptides. The secondary outcome was the total IgG antibody responses to the *Aedes* N-term 34 kDa salivary peptides. Adverse events were assessed at every visit. Prespecified adverse events that would have required withdrawing the participant from the study included skin reactions greater than 30 mm in diameter, ecchymosis, vesicle, blister, bullae, Skeeter syndrome and systemic symptoms (generalized urticaria, angioedema and anaphylaxis).

### Statistical analysis

The change in individual antibody responses (as a continuous outcome and binary response) over time, and in response to mosquito biting exposures, were estimated according to the protocol using generalized linear mixed-effects models. Given we observed high levels of serial correlation in antibodies within individuals over time, generalised estimating equations (GEE) with robust standard errors were fitted to the longitudinal antibody data to provide robust inference of the estimated effects of the exposure and post-exposure periods on antibody levels.

The analysis estimates the change in mean antibody levels (log_2_ transformed) associated with the intervention (dummy indicators for biting exposure and post-exposure periods compared to the baseline), includes a linear term for time to measure per day changes in antibodies independent of the biting interventions, and is adjusted for age and sex. The potential effect modification of the intervention group was explored by estimating interaction terms between the intervention period and intervention group (i.e. mosquito biting species and dose). Given we observed waning correlation in within-individual antibody levels over time, a 1^st^ order autoregressive working correlation structure was used in the GEE analyses to provide correct inference given the dependencies in the data (repeated antibody measurements). The statistical analyses were performed using STATA v18 and plots were generated in R.

## Results

Between January 21, 2021, and May 10, 2022, 248 volunteers were screened for eligibility, 212 were enrolled, 210 contributed at least one sample and 204 were challenged at least once with mosquito bites (Figure 1). The study was interrupted on July 15, 2021 because of a COVID-19 outbreak at the study site and was resumed on November 3, 2021. At the time of study interruption, 55 participants had not completed the study; eleven participants who had attended 19/20 visits were not replaced and 44 participants who had attended 17 visits or less were replaced with new participants when the study resumed. A total of seven participants had incomplete follow-up because of an adverse event (detailed in Appendix Table 3). All participant data, excluding four individuals who only contributed one sample and were not challenged with mosquito bites, were included in the intention-to-treat analysis (n=206, Figure 1). Only 18 participants reported traveling to rural areas outside Mae Sot city during the study. All participants travelled only once, the median duration of overnight stay was four nights (IQR 2-5·5, range 0-15), only four (22%) travelled during the rainy season, and 14 participants (78%) reported using personal protection with mosquito-bed nets and/or skin repellents during those trips.

**Figure 1.**
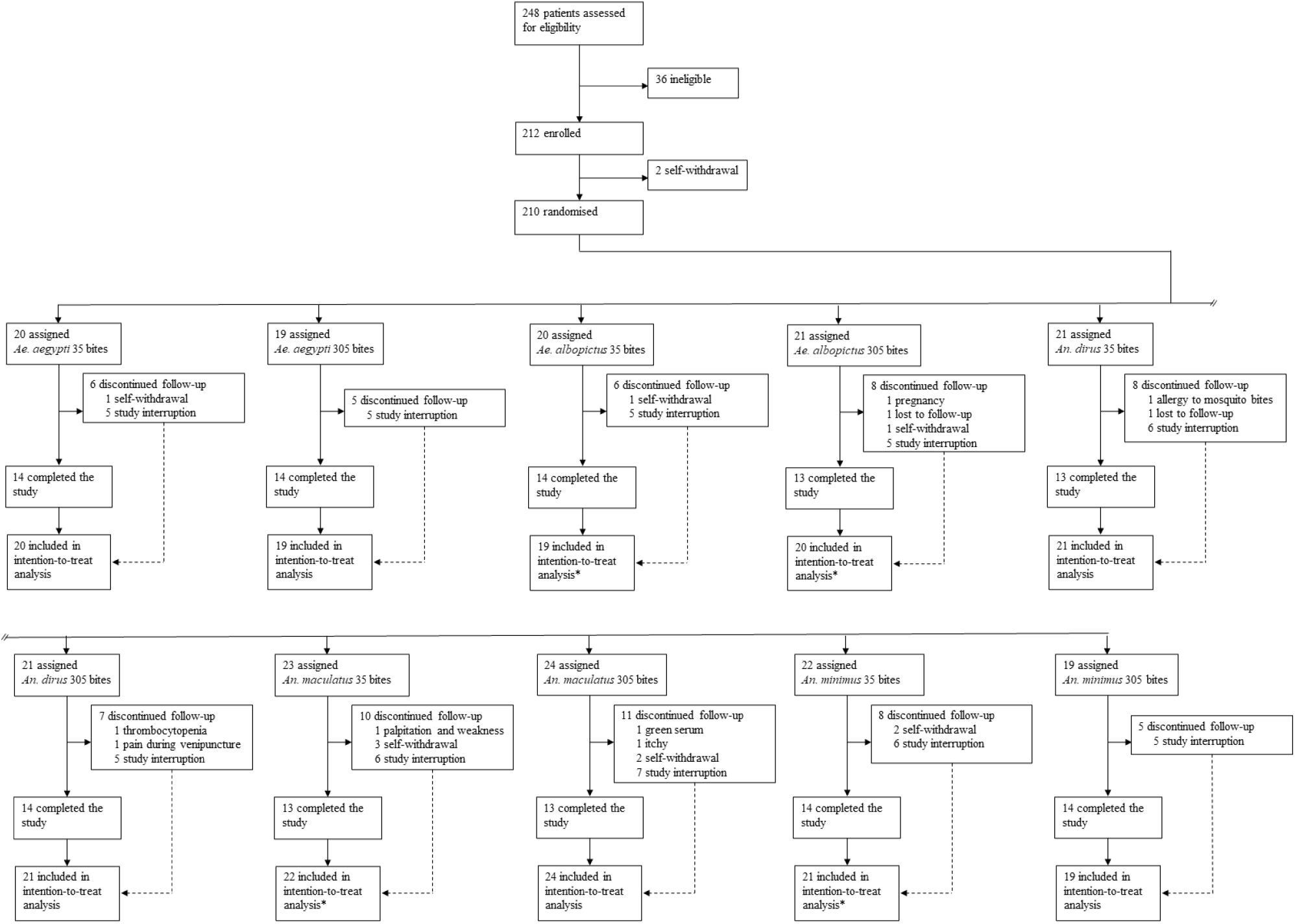
Flow diagram of trial design.

Participants’ baseline characteristics are shown in Table 1. Overall, 115 (54%) participants were male and the median age was 30 years (inter-quartile range [IQR] 25-39·2). Participants had relatively high levels and seroprevalence of antibodies against *Anopheles* and *Aedes* salivary antigens (ranging from 50-78% across antigens) at baseline (prior to any challenge) compared to unexposed participants from Melbourne, Australia who had low levels of antibodies and were all seronegative (Figure 2). Baseline antibody levels were also found to vary according to enrolment date, with higher levels of antibodies observed prior to the interruption of the study (February – June 2021 compared to November 2021 – May 2022) (Appendix Figure 2).

**Table 1.** Baseline characteristics of the cohort.

|  | AEG_35 (n=20) | AEG_305 (n=19) | ALB_35 (n=20) | ALB_305 (n=21) | DIR_35 (n=21) | DIR_305 (n=21) | MAC_35 (n=23) | MAC_305 (n=24) | MIN_35 (n=22) | MIN_305 (n=19) |
| --- | --- | --- | --- | --- | --- | --- | --- | --- | --- | --- |
| **Sex** |  |  |  |  |  |  |  |  |  |  |
| *Male* | 12 (60%) | 10 (53%) | 9 (45%) | 9 (43%) | 8 (38%) | 15 (71%) | 12 (52%) | 16 (67%) | 14 (64%) | 9 (47%) |
| *Female* | 8 (40%) | 9 (47%) | 11 (55%) | 12 (57%) | 13 (62%) | 6 (29%) | 11 (48%) | 8 (33%) | 8 (36%) | 10 (53%) |
| **Age (years)** | 31·5 (25-41) | 30 (27-34·5) | 31·5 (26·2-37·2) | 29 (27-43) | 28 (24-32) | 31 (24-47) | 26 (22-33) | 33·5 (27·8-44·2) | 35·5 (25·5-39·8) | 30 (26-37) |
| **gSG6-P1** |  |  |  |  |  |  |  |  |  |  |
| *Levels (OD)* | 2·3 (1·54-2·51) | 2·11 (1·66-2·44) | 2·14 (1·5-2·43) | 2·05 (1·43-2·49) | 2·14 (1·48-2·47) | 2·26 (1·75-2·49) | 1·99 (1·58-2·51) | 2·14 (1·39-2·48) | 1·96 (1·59-2·5) | 1·83 (1·29-2·46) |
| *Seroprevalence* | 46/59 (78%) | 46/57 (80·7%) | 46/58 (79·3%) | 46/61 (75·4%) | 50/63 (79·4%) | 52/62 (83·9%) | 55/67 (82·1%) | 53/72 (73·6%) | 51/64 (79·7%) | 38/57 (66·7%) |
| **minSG6-P1** |  |  |  |  |  |  |  |  |  |  |
| *Levels (OD)* | 0·84 (0·4-1·34) | 0·72 (0·51-1·15) | 0·71 (0·44-1·07) | 0·69 (0·44-1·08) | 0·67 (0·37-1·34) | 0·75 (0·49-1·19) | 0·71 (0·4-1·08) | 0·65 (0·42-0·97) | 0·64 (0·4-1·02) | 0·55 (0·33-1·03) |
| *Seroprevalence* | 33/59 (55·9%) | 31/57 (54·4%) | 31/58 (53·4%) | 31/61 (50·8%) | 31/63 (49·2%) | 35/62 (56·5%) | 36/67 (53·7%) | 34/72 (47·2%) | 31/64 (48·4%) | 21/57 (36·8%) |
| **macSG6-P1** |  |  |  |  |  |  |  |  |  |  |
| *Levels (OD)* | 0·81 (0·5-1·21) | 0·64 (0·43-0·85) | 0·6 (0·4-0·94) | 0·59 (0·4-1·16) | 0·67 (0·47-1·43) | 0·78 (0·43-1·07) | 0·67 (0·47-1·22) | 0·57 (0·45-1·07) | 0·57 (0·4-1·01) | 0·53 (0·38-0·86) |
| *Seroprevalence* | 41/59 (69·5%) | 35/57 (61·4%) | 31/58 (53·4%) | 31/61 (50·8%) | 41/63 (65·1%) | 40/62 (64·5%) | 41/67 (61·2%) | 40/72 (55·6%) | 34/64 (53·1%) | 27/57 (47·4%) |
| **dirSG6-P1** |  |  |  |  |  |  |  |  |  |  |
| *Levels (OD)* | 1·3 (0·63-1·79) | 1·07 (0·72-1·51) | 0·96 (0·63-1·51) | 1·11 (0·68-1·97) | 1·12 (0·63-1·97) | 1·16 (0·69-2·13) | 1·05 (0·72-1·73) | 0·98 (0·64-1·66) | 0·99 (0·61-1·77) | 0·91 (0·56-1·54) |
| *Seroprevalence* | 40/59 (67·8%) | 42/57 (73·7%) | 35/58 (60·3%) | 43/61 (70·5%) | 44/63 (69·8%) | 44/62 (71%) | 50/67 (74·6%) | 45/72 (62·5%) | 43/64 (67·2%) | 32/57 (56·1%) |
| **aeg34kDa** |  |  |  |  |  |  |  |  |  |  |
| *Levels (OD)* | 1·61 (1·06-2·13) | 1·64 (1·22-2·12) | 1·43 (0·9-1·97) | 1·65 (0·88-2·08) | 1·88 (1·15-2·18) | 1·70 (1·1-2·05) | 1·47 (1·05-2·1) | 1·73 (0·96-2·12) | 1·55 (0·82-2·07) | 1·51 (1·12-2·03) |
| *Seroprevalence* | 45/59 (76·3 %) | 46/57 (80·7 %) | 45/58 (77·6 %) | 46/61 (75·4 %) | 54/63 (85·7 %) | 50/62 (80·6 %) | 57/67 (85·1 %) | 54/72 (75 %) | 45/64 (70·3 %) | 47/57 (82·5 %) |
| **alb34kDa** |  |  |  |  |  |  |  |  |  |  |
| *Levels (OD)* | 1·56 (0·84-2·19) | 1·5 (0·87-2·12) | 1·28 (0·88-1·95) | 1·52 (0·86-2·19) | 1·76 (1·03-2·18) | 1·68 (1·02-2·02) | 1·35 (0·89-2·06) | 1·65 (0·91-2·02) | 1·38 (0·77-2·1) | 1·45 (0·95-1·97) |
| *Seroprevalence* | 44/59 (74·6 %) | 43/57 (75·4 %) | 46/58 (79·3 %) | 46/61 (75·4 %) | 52/63 (82·5 %) | 49/62 (79 %) | 55/67 (82·1 %) | 55/72 (76·4 %) | 46/64 (71·9 %) | 44/57 (77·2 %) |

Data are n (%), median (IQR), or n/N (%) for all randomised participants (n=210). Antibody levels and seroprevalence were calculated using the 3 baseline samples collated by participant. OD – optical density units

**Figure 2.**
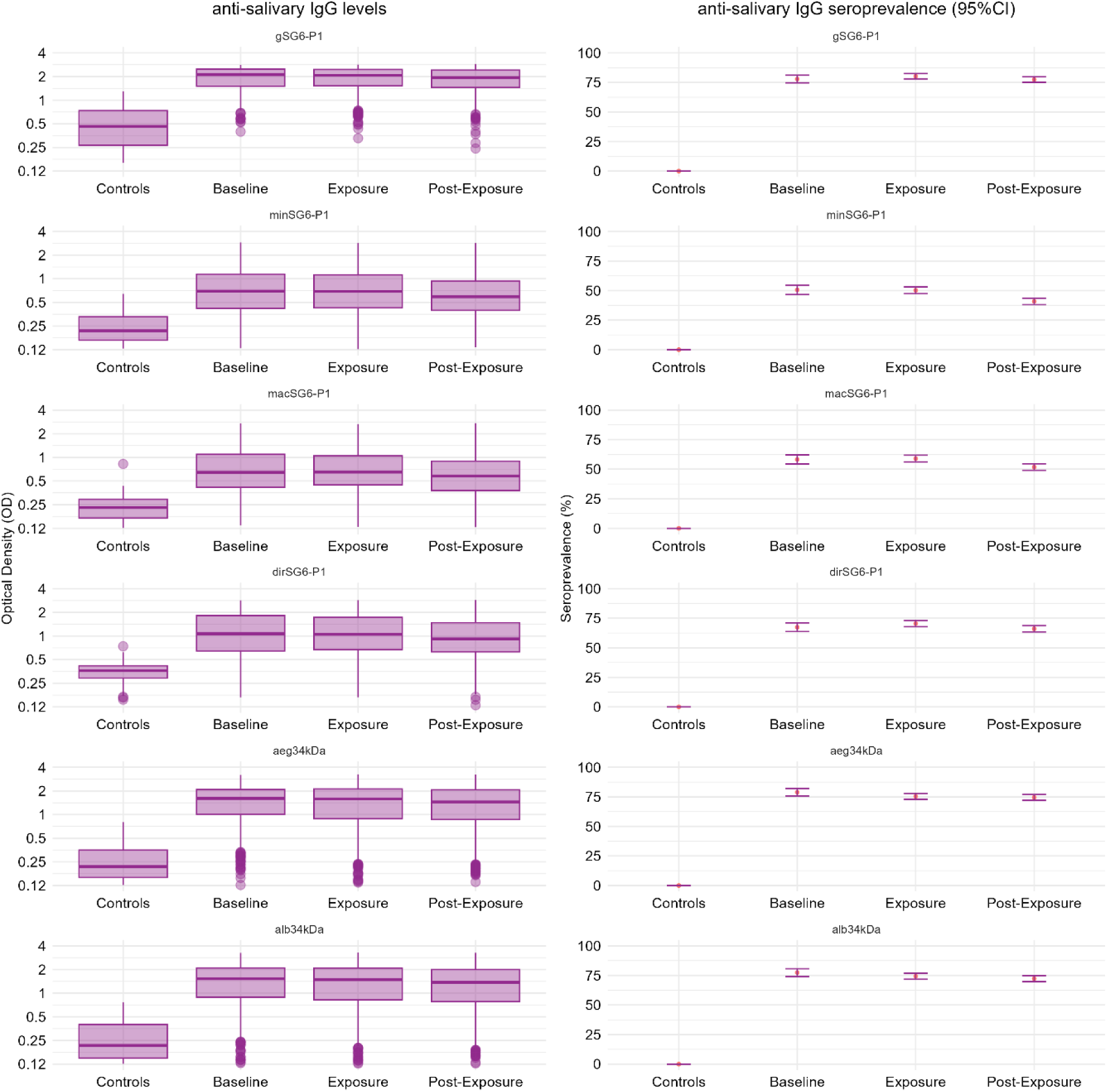
Antibody response by period, compared to Melbourne controls.

Antibody responses against *Anopheles* salivary SG6-P1 antigens were highly dynamic over time, both within and between individuals (observed individual- and population-level responses by intervention group are presented in Figure 3, and Appendix Figures 3-7). Analyses investigating the overall effect of mosquito biting exposure determined that IgG antibodies to *Anopheles* SG6-P1 salivary antigens decayed over time but were associated with boosts in antibody levels during the exposure and post-exposure periods (Appendix Table 6). The magnitude and confidence intervals associated with these estimates varied according to the antigen investigated and are detailed in Appendix. The rate of antibody decay following biting exposure (i.e. half-life for the post-exposure period) was found to range from 193 (95%CI: 108 - 278) to 421 (95%CI: 155 - 688) days depending on the SG6-P1 ortholog (Figure 4). The species- and dose-specific effects of mosquito biting exposure on the anti-salivary antibody response were determined. The largest boosts in antibody levels and seroprevalence during the exposure and post-exposure periods were observed amongst the individuals exposed to a high dose (305 bites over 6 weeks) of *An. minimus* bites. For example, anti-gSG6-P1 IgG antibody levels for the high-dose *An. minimus* group were boosted 1.14-fold (geometric mean ratio) (95%CI: 1.03-1.26) and 1.18-fold (95%CI: 1.05-1.33) during the exposure and post-exposure periods, respectively (relative to baseline), with minimal or no boosting in IgG levels observed in other groups (Figure 5). Similarly, the largest increases in the odds of anti-gSG6-P1 IgG seropositivity during the exposure (OR: 2.94, 95%CI: 2.71-5.05) and post-exposure (OR: 5.54, 95%CI: 2.73-11.24) periods were observed for the high-dose *An. minimus* intervention group (Figure 6). Similar associations were observed across SG6-P1 orthologs.

**Figure 3.**
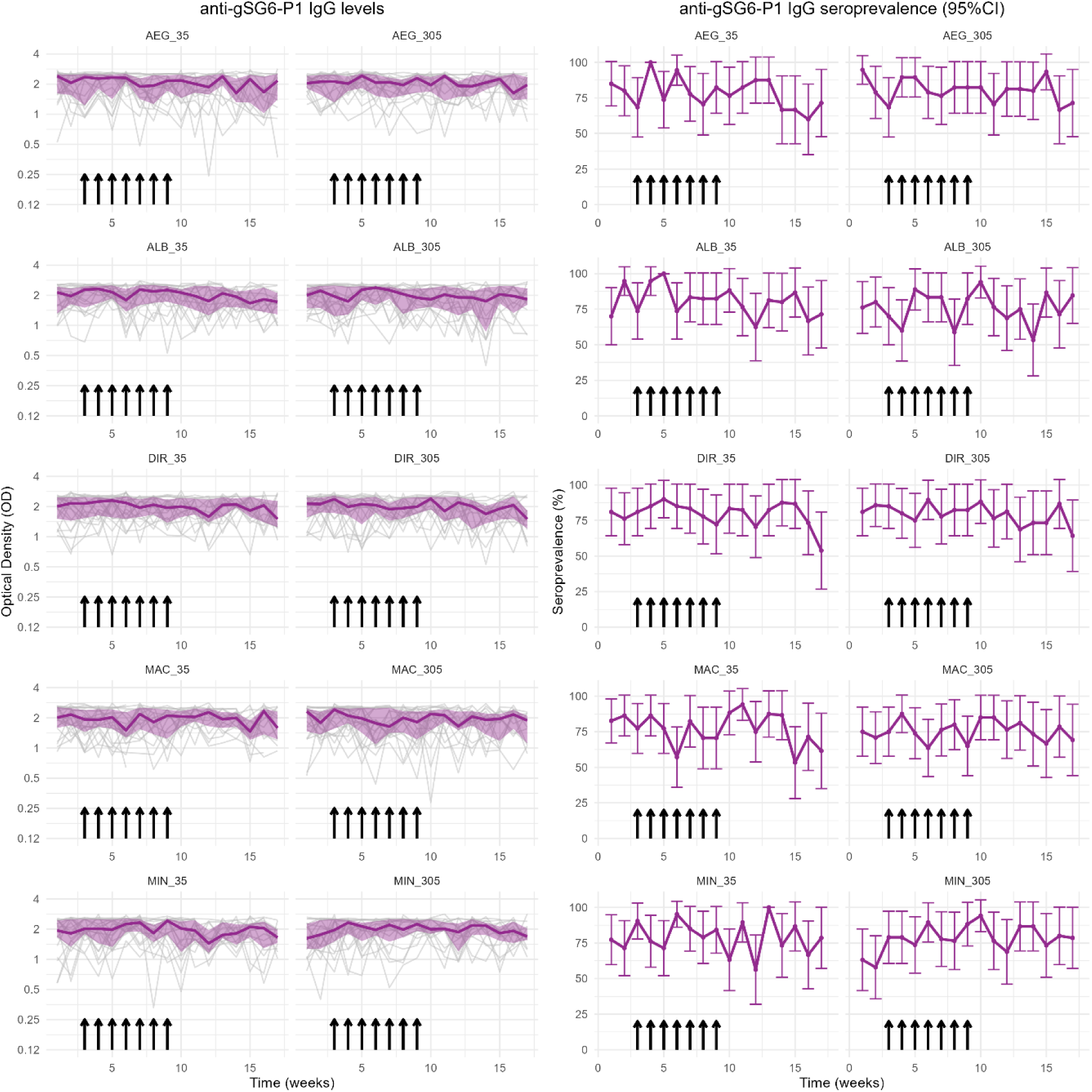
Individual- and population-level antibody responses against gSG6-P1 over time, by intervention group.

**Figure 4.**
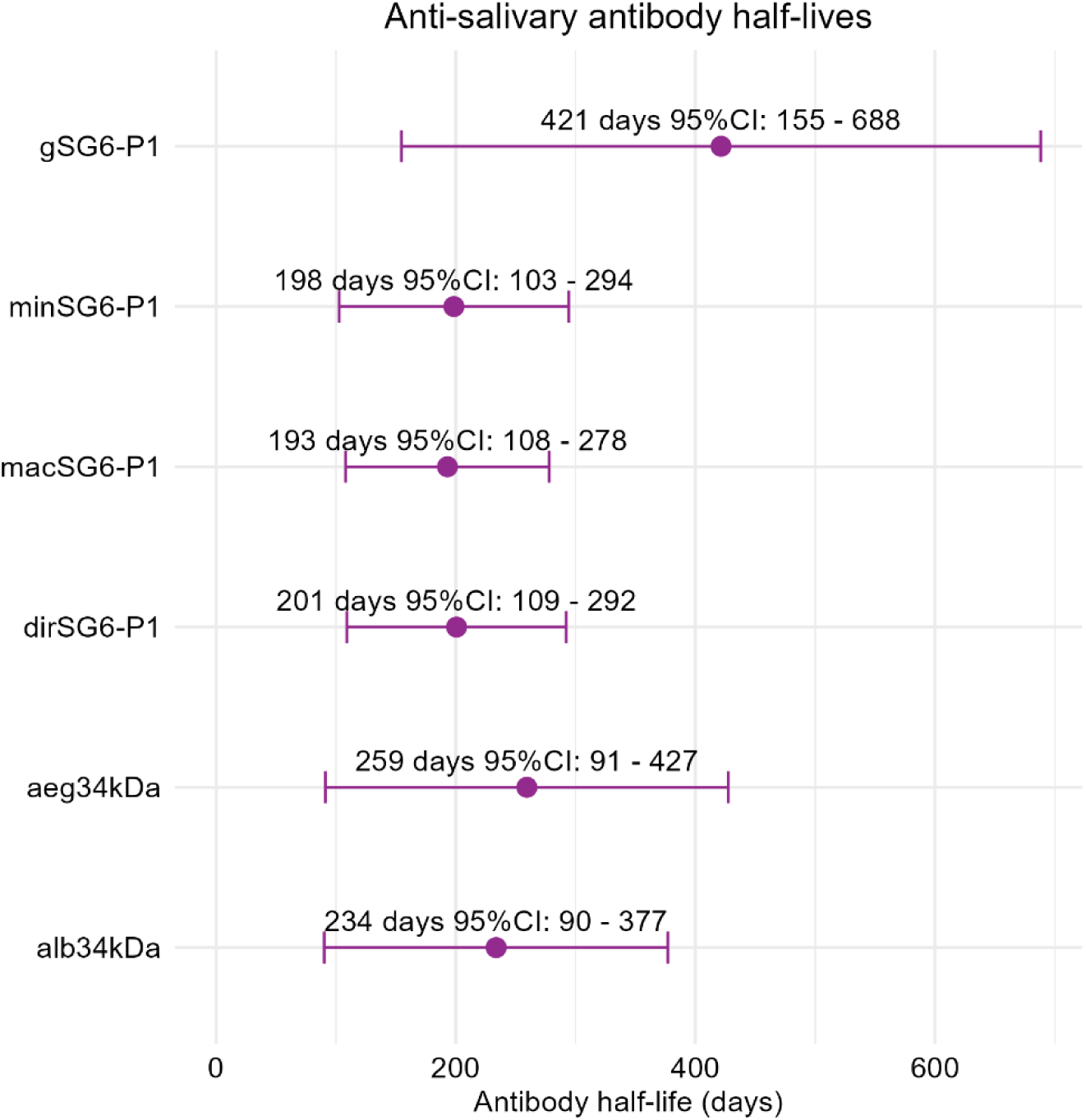
Anti-salivary antibody rates of decay.

**Figure 5.**
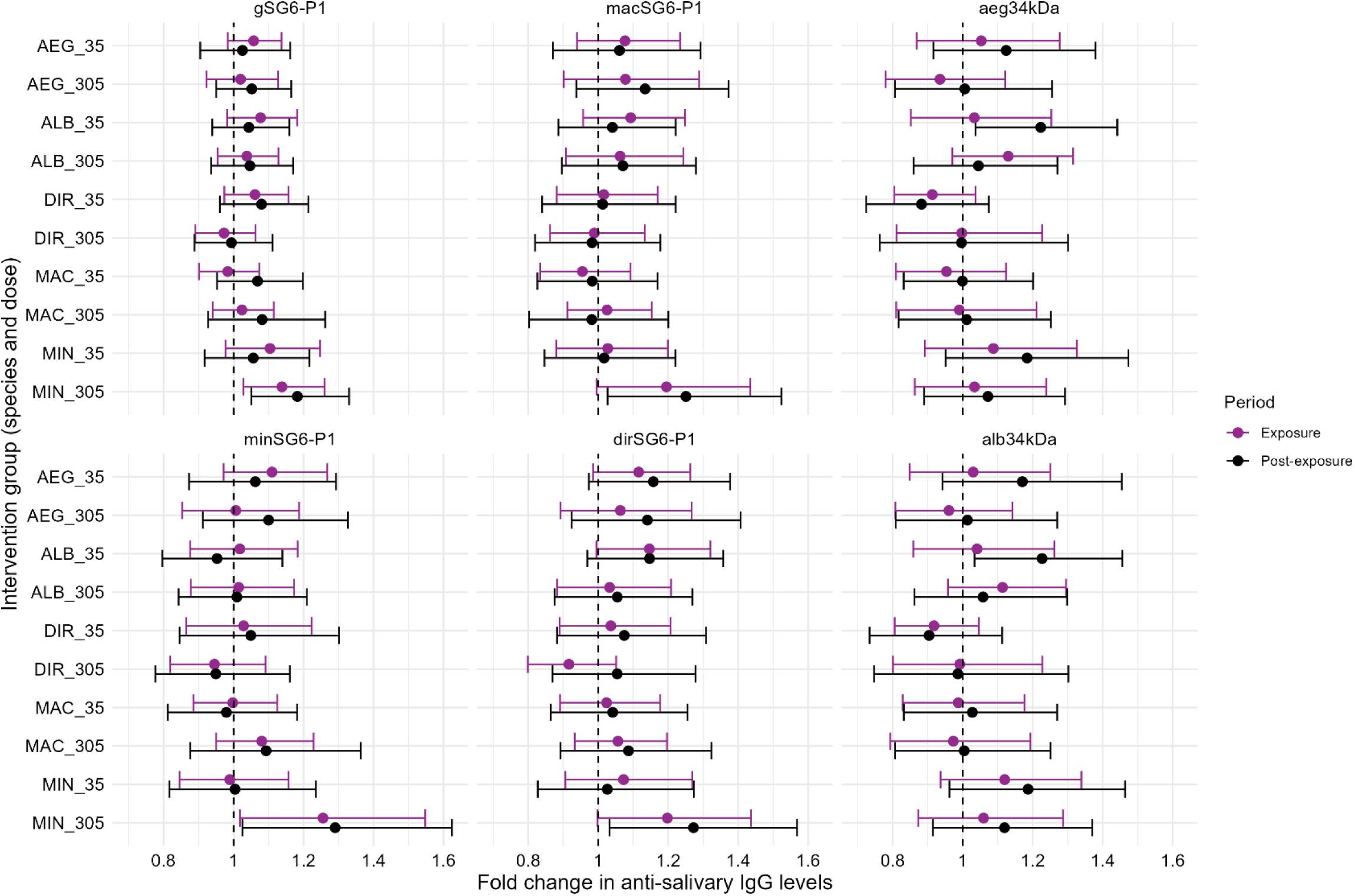
Fold changes in anti-salivary antibody levels associated with biting exposures for each of the intervention groups.

**Figure 6.**
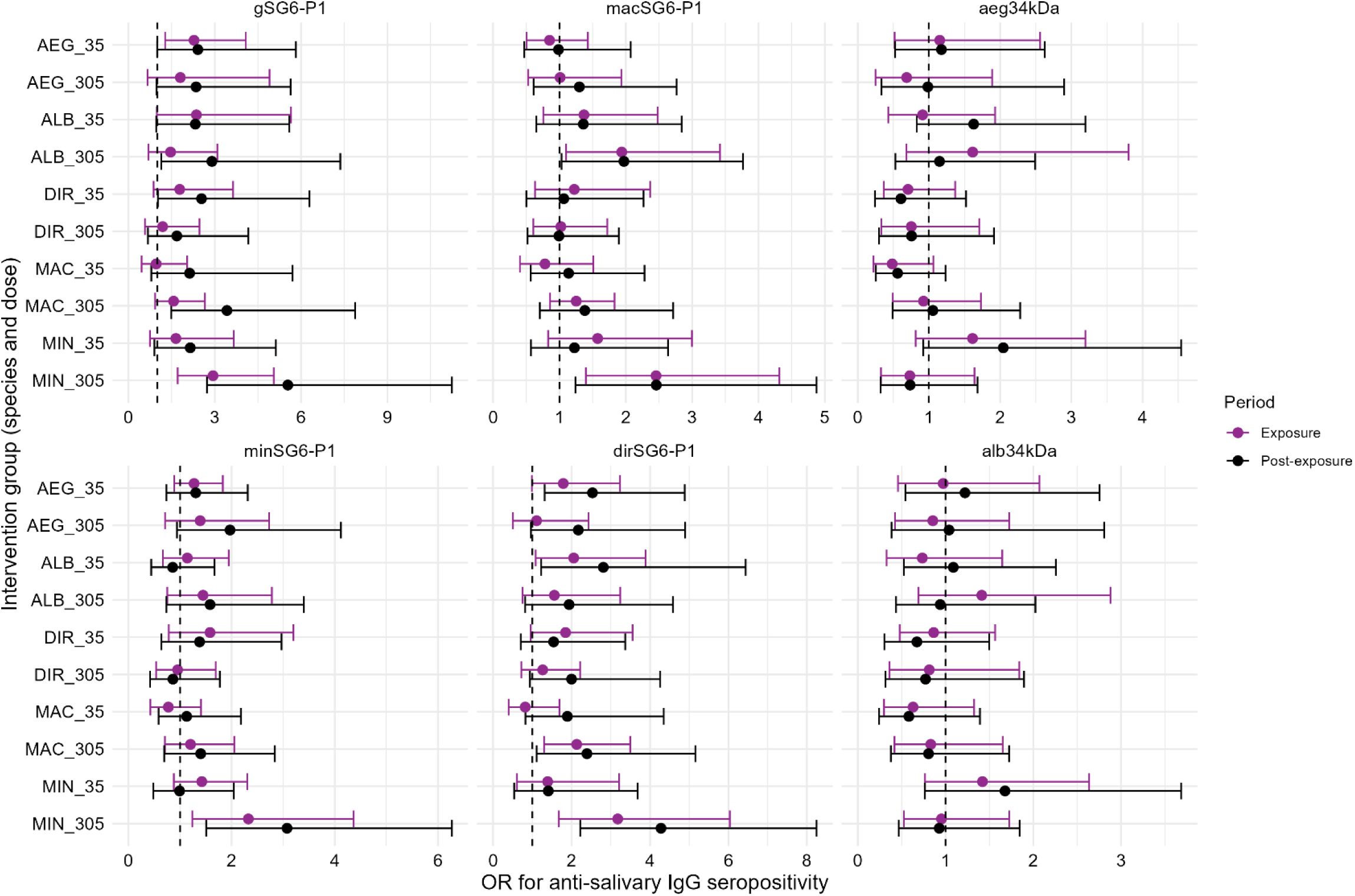
Odds ratios for anti-salivary antibody seroprevalence associated with biting exposures for each of the intervention groups.

Antibody responses to aeg34kDa and alb34kDa salivary antigens were also dynamic over time, and were found to have respective half-lives of 259 (95%CI: 90 – 427) and 234 (95%CI: 90 - 377) days following biting exposure (Figure 4). The largest boosts in antibody levels were observed amongst the 35 *Ae. albopictus* biting group: in this group, anti-aeg34kDa and anti-alb34kDa IgG levels increased 1.22-fold (95%CI: 1.04-1.44) and 1.22-fold (95%CI: 1.03-1.46) in the post-exposure period relative to baseline, respectively (Figure 5). Similar boosts were observed amongst those exposed to 35 bites of *Ae. aegypti,* however, the confidence intervals associated with these estimates were wide (aeg34kDa: 1.12-fold, 95%CI: 0.92-1.38; alb34kDa: 1.17, 95%CI: 0.94-1.45). Mosquito biting exposures were not associated with significant increases in anti-*Aedes* salivary antibody levels in the other intervention groups.

## Discussion

In this world-first trial of controlled exposure to vector bites, total IgG responses to *Anopheles* SG6-P1 and *Aedes* N-term 34kDa salivary peptides were assessed in volunteers bitten by mosquitos of laboratory-adapted colonies of *An. dirus*, *An. maculatus*, *An. minimus* (the main malaria vectors in the Greater Mekong Subregion), *Ae. aegypti*, and *Ae. albopictus* (the main dengue vectors worldwide). Most participants had detectable antibodies prior to mosquito biting challenges and total IgG responses to SG6-P1 peptides were boosted during and after exposures to 305 bites of *An. minimus* split in 7 challenges over 6 weeks, but not after exposures to smaller numbers of bites or bites of other vector species. Similarly, total IgG antibody responses to 34kDa peptides were boosted following exposure to 35 total bites of *Ae. albopictus* but were not associated with significant boosts in antibody responses in the other groups. Overall, the increases in antibody titres were relatively small and anti-salivary antibodies decayed slowly after cession of exposure.

This study identified that significant boosts in antibodies against gSG6-P1 were only observed for the group exposed to a large number of bites from *An. minimus*, whose SG6-P1 peptide sequence shares the greatest identity to gSG6-P1 (87%^10^). This suggests that the level of identity of the SG6 ortholog (injected during bite exposure from a non-*An. gambiae* species) to gSG6-P1 may be important in determining the degree of antibody cross-reactivity. This has important implications for the use of gSG6 outside of Africa, as low cross-reactivity may reduce its sensitivity and limit its application as genus-level biomarker of exposure to the bites of all relevant *Anopheles* mosquito species. Antibody responses to the SG6-P1 orthologs were similar over time and in response to mosquito biting exposures, with boosting upon biting exposure consistently identified only amongst the *An. minimus* high-exposure arm (305 bites per person). The weaker non-significant boost in antibody levels across the SG6-P1 antigens for the *An. maculatus* and *An. dirus* exposure groups may be a result of their lower sequence identity to gSG6-P1 (78% and 48% respectively^10^) potentially resulting in less immunogenic epitopes from the P1 region being present in these novel antigens. Additional novel *Anopheles* salivary antigens corresponding to alternate regions of the SG6 protein (which bioinformatic analysis has predicted higher immunogenicity in the terminal region, Appendix Figure 1) or one of dozens of other identified *Anopheles*-specific salivary proteins^10^ could readily be investigated and validated as sensitive biomarkers of species-specific exposure in this cohort.

Study participants were found to have relatively high (yet variable) levels of anti-SG6-P1 and anti-34kDa antibodies at baseline, particularly compared to unexposed controls from Melbourne, Australia. This suggests that these antibodies recognise prior biting exposure to bites of *Anopheles* and *Aedes* mosquitos. These baseline antibody responses were ideal to mimic people naturally exposed to vector bites in endemic areas, as are the participants enrolled in trials of vector control and serosurveillance studies. As we excluded participants who had travelled to rural areas in the past 12 months, this finding is consistent with the slow decay rate of anti-salivary antibodies reported in this assessment. The anti-gSG6-P1 IgG half-life estimate reported in this assessment is comparable to the only other published estimate (424 days^24^ which was determined without detailed knowledge of biting exposure but instead time since malaria infection). These long antibody half-lives challenge previous claims that total IgG responses to SG6-P1 peptides decay rapidly following interrupted exposure to mosquito bites^25^. This result has important implications for how these anti-salivary antibodies can be used in future studies. For example, they may be better suited to investigating longer-term changes in transmission dynamics, and studies employing them as outcomes in vector control trials will need to allow for sufficient follow up to evaluate the delayed effects of the intervention so that its effectiveness is not underestimated due to the persistence of antibodies after biting exposure is reduced. As different antibody isotypes and subclasses have more dynamic responses than others^26^, additional work is on-going to characterise their distinct longitudinal profiles within this cohort.

The high levels and seroprevalence of IgG antibodies at baseline (prior to controlled biting) may have contributed to the relatively small magnitude of antibody boosting associated with biting exposure observed in this trial, suggesting the development of an immune tolerance after repeated biting exposures. Evidence for this tolerance in previous studies is conflicted, reporting both positive and negative associations between anti-salivary antibodies and age^12,27^. Alternatively, minimal boosting may be a result of the relatively small antigenic stimulus of mosquito salivary proteins injected during a bite and may suggest that individuals may need to be exposed to greater numbers of bites to register larger boosts in antibody levels. As some entomological surveys from the Greater Mekong Subregion report biting rates of over 600 bites per person per month^3^, these salivary antibody biomarkers may still be applicable to quantify variation in biting rates throughout the region. Additional investigations of the sensitivity of these antibodies to detect differing levels of biting exposures are warranted.

Due to the complexity of quantifying an individual’s natural mosquito biting exposures, this trial uses a human challenge framework to control participants’ biting exposures and characterise the longitudinal dynamics of the anti-salivary antibody responses. This novel approach is a key strength of this study, which provides a valuable framework for future research striving to develop biomarkers of recent exposures to vector bites. However, there are several limitations to acknowledge. We used a factorial design because the trial was exploratory and the sample size per group was small. Therefore, the study was not powered to detect small-sized effects. Due to the ubiquitous nature of *Aedes* mosquitoes, unobserved exposures to *Aedes* bites not related to the study challenges is likely to have occurred, potentially confounding the estimation of anti-34kDa antibody boosting and decay rates. While we attempted to control for this by providing participants with topical repellent, repellent use was not directly observed or reported. The significant variation in the baseline antibody responses observed across the enrolment dates was difficult to explain, but may have been caused by seasonality, past history of exposure to *Anopheles* bites, or travel restrictions during the COVID-19 pandemic. Moreover, baseline IgG responses to SG6-P1 peptides were lower in the groups of participants challenged with 305 bites of *An. minimus* which may have confounded the estimated effect of biting exposure on antibody levels within this group. We used five-to-seven-day-old nulliparous mosquitos of laboratory-adapted colonies, and the expression of SG6 was not characterized. Previous studies have reported variation of mosquito salivary proteome with mosquito age^28^, infection^29^ and resistance^30^. Hence, the proteome of the mosquitos used in this study may be different than that of wild mosquitos biting humans in natural settings. Although a more detailed characterisation of salivary gland proteome was out of the scope of this study, this should be the focus of future research. Only SG6-P1 and N-term 34 kDa peptides were tested. Although synthetic peptides are useful for high-throughput screening at scale, they do not capture conformational epitopes and epitopes of other types of biomolecules (e.g., glycoproteins) that may be important targets of antibody responses to mosquito salivary components.

Future research should strive to screen additional targets, including whole saliva protein extract, salivary proteins in their native form, and synthetic peptides. Mosquito species diversity is particularly high in this region and many relevant vector species were not assessed. Colonization of mosquitos is challenging and many vectors have not been colonized yet. Additional research striving to establish colonies for relevant mosquito vector species should be encouraged for inclusion in future trials of controlled mosquito biting exposure. Furthermore, this vector salivary biomarker approach could readily be extended to other medically important arthropods and combined with pathogen serology as an interesting approach to surveillance of vector-borne diseases, but additional research is needed before these tools can be implemented programmatically^23^.

In conclusion, our study demonstrated small boosts in antibody responses directed against mosquito salivary antigens following controlled exposure to mosquito bites in a population with high levels of antibodies during baseline. Additional effort is needed to develop reliable biomarkers of recent exposures to vector bites.

## Data Availability

All data produced in the present study are available upon reasonable request to the authors

## Acknowledgement

We are very grateful to the volunteers who participated in this study. We thank the staff of the Entomology, Laboratory, Medical, and Data Management Departments of the Shoklo Malaria Research Unit for their help with collection, processing, and management of the samples and data included in this study. We thank the Clinical Trial Support Group of the Mahidol-Oxford Research Unit for their support to this trial. The Shoklo Malaria Research Unit is part of the Mahidol-Oxford Research Unit, supported by Wellcome, U.K. (#220211). This research was funded by Wellcome. A CC BY or equivalent license is applied to the author accepted manuscript arising from this submission, in accordance with the grant’s open access conditions. Antibody data generation was supported by the Australian National Health and Medical Research Council (NHMRC) eASIA Joint Research Program grant (#2009656) awarded to EAK, PAA, KO, JAS and FJIF. JAS and FJIF are funded by NHMRC Leadership Investigator Grants (# 1196068 and 2017485). The Burnet Institute is supported by a Victorian State Government Operational Infrastructure Support grant. The funders had no role in the decision to publish this manuscript.

## Figure list

Appendix, Figure 1: Peptide design

Appendix; Figure 2: Antibody data by enrolment month

Appendix; Figure 3. Individual- and population-level antibody responses against minSG6-P1 over time, by intervention group.

Appendix; Figure 4. Individual- and population-level antibody responses against macSG6-P1 over time, by intervention group.

Appendix; Figure 5. Individual- and population-level antibody responses against dirSG6-P1 over time, by intervention group.

Appendix; Figure 6. Individual- and population-level antibody responses against aeg34kDa over time, by intervention group.

Appendix; Figure 7. Individual- and population-level antibody responses against alb34kDa over time, by intervention group.

## Table list

Appendix Table 1. Pairwise comparison of SG6-P1 sequences across SG6 orthologs from the Southeast Asian malaria vector species and N-term 34kDa sequence across 34kDa orthologs of global dengue vectors.

Appendix Table 2. Adverse events

Appendix Table 3. Observed values of total IgG antibody responses by intervention group and follow-up period.

Appendix Table 4. Effect of mosquito biting exposure period (across all intervention groups) on anti-salivary antibody levels.

Appendix Table 5. Time-dependent effect of mosquito biting exposure period (across all intervention groups) on boosting and decay of anti-salivary antibodies.

Table 6. Effect of mosquito biting exposure period, modified by intervention group (species and dose), on anti-salivary antibody levels.

Table 7. Effect of mosquito biting exposure, modified by intervention group (species and dose), on anti-salivary antibody seroprevalence.

