## Appendix for "Assessment of antibody responses to *Anopheles* SG6-P1 and *Aedes* N-term 34kDa salivary peptides: a human-challenge trial of controlled exposures to vector bites"

#### Supplementrary Methodology

Spectraplates (Perkin Elmer) were coated with 0.5µg/mL of mosquito salivary peptide (Genscript) resuspended in autoclaved MilliQ water and diluted in phosphate buffered saline (PBS), and incubated for 3 hours at 37ºC. Plates were washed and blocked for one hour at 37ºC with blocking buffer (Pierce, Thermo Scientific USA). After a subsequent wash step, sera were added at desired concentrations (1:1400 for all SG6-P1 antigens; 1:2800 for aeg34kDa; 1:3600 for alb34kDa) diluted in 10% blocking buffer with PBS and incubated overnight at 4ºC. Following sera incubation, plates were washed and secondary antibody added. To detect human IgG, horseradish peroxidase- (HRP) conjugated goat anti-human IgG (Millipore) was used at a 1:500 dilution. Plates were incubated at 37ºC for 1·5 hours and then washed. ABTS (2,2'-azino-bis(3-ethylbenzothiazoline-6-sulfonic acid)) substrate was added to each well, covered and left to develop at room temperature, then stopped with 1% sodium dodecyl sulphate, and the optical density (OD) was read in a spectrophotometer at 405 nm. Seropositivity for each peptide was defined as having an OD greater than the mean plus three standard deviations of the unexposed Melbourne controls (n=8).

### Supplementrary Figures

A

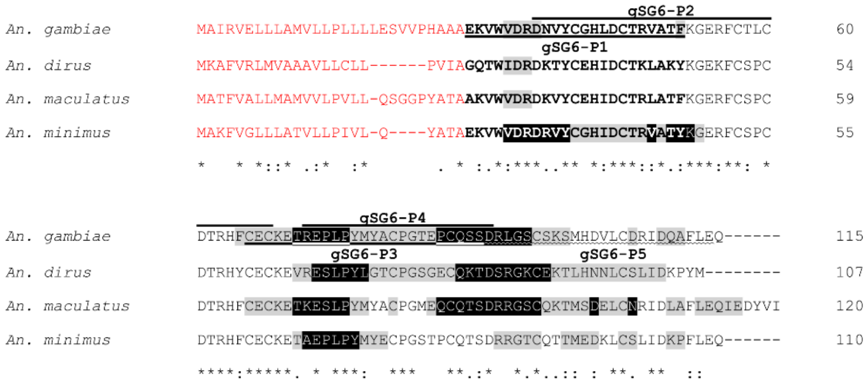

B

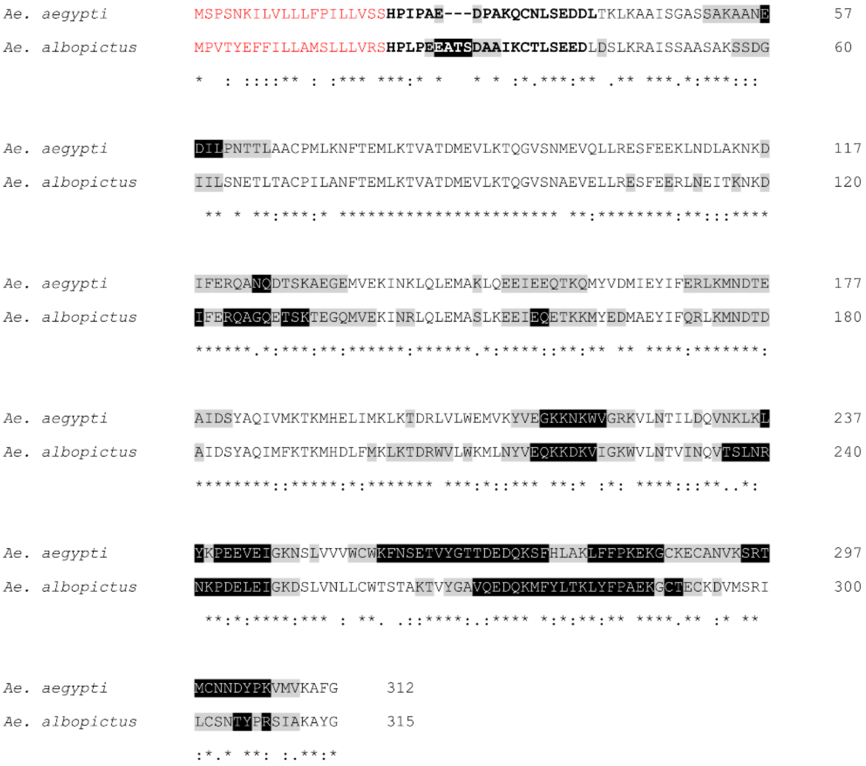

**Figure 1. Alignment of salivary gland protein 6 (SG6) from Southeast Asian *Anopheles* species (A) and 34kDa salivary protein from global dengue vectors (B).** Publicly available sequences in VectorBase repository and cured by others were downloaded from the Additional File 19 in Arca *et al.* paper (BMC Genomics, 2017). gSG6-P1, gSG6-P2, gSG6-P3, gSG6-P4, gSG6-P5 are the candidate peptides identified by Poinsignon *et al.* (PLOS One, 2008). Signal peptide identified with SignalP 6.0 (Nielsen *et al.* Methods Mol biol, 2024) are showed in red and were removed prior to epitope prediction with Prediction 3.0 (Clifford *et al.* Protein Sci, 2022). The top 20% and 50% most likely B-cell epitope predictions specifying sequential smoothing (rolling mean) in the analysis are highlighted in black and in grey respectively. The residues in bold show the peptides tested in this study. (*) positions that have a single and fully conserved residue; (:) conservation between groups of strongly similar properties with a score greater than .5 on the PAM 250 matrix, (.) conservation between groups of weakly similar properties with a score less than or equal to .5 on the PAM 250 matrix.

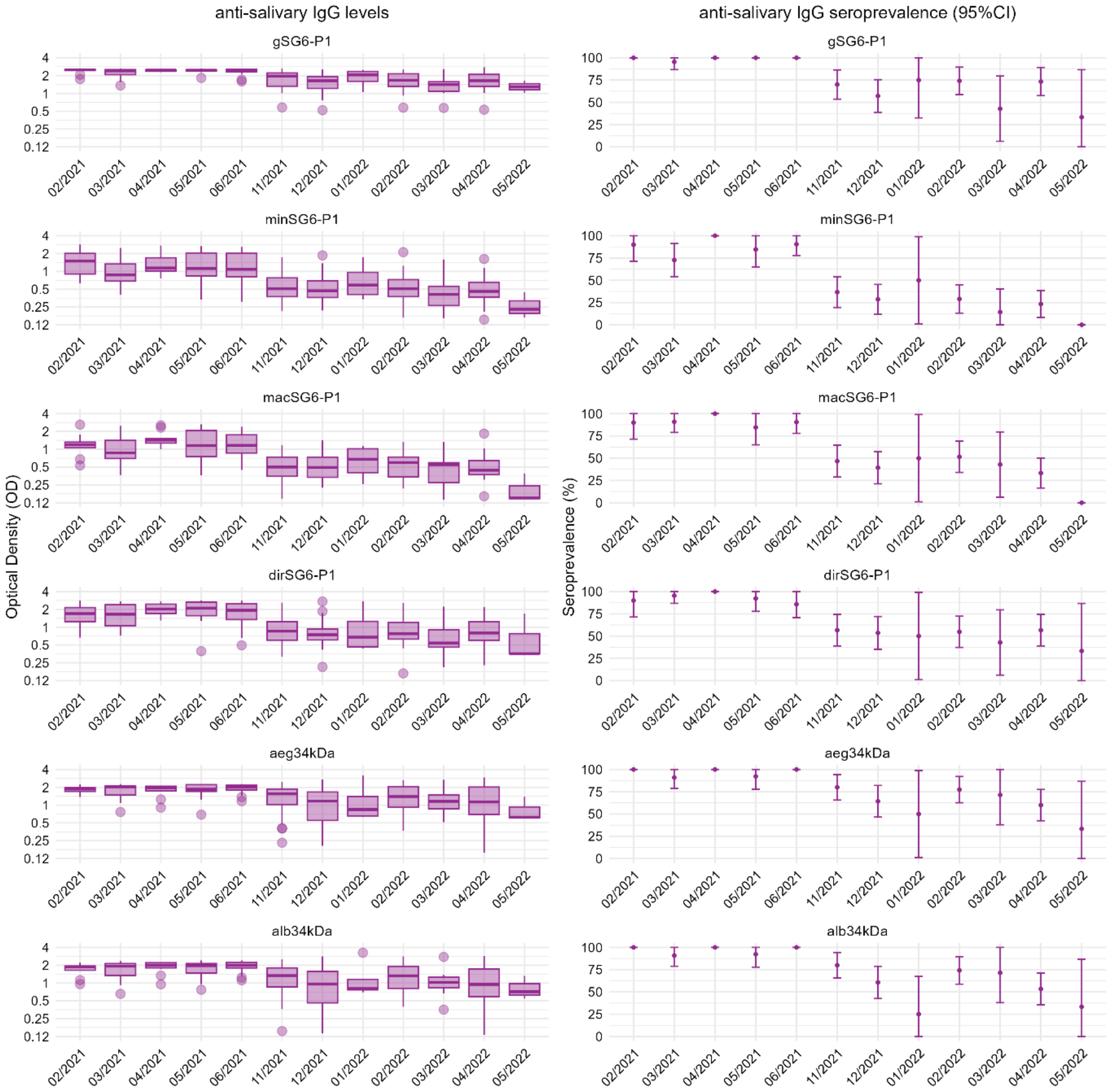

**Figure 2. Anti-salivary antibody data at first baseline visit by enrolment month.** Box plots on the left show the median and interquartile range of antibody levels (OD) and bar charts on the right show the seroprevalence (95% Confidence Interval (CI)) of antibody responses against each salivary peptide at participants first baseline visit (prior to any exposure) by enrolment month.

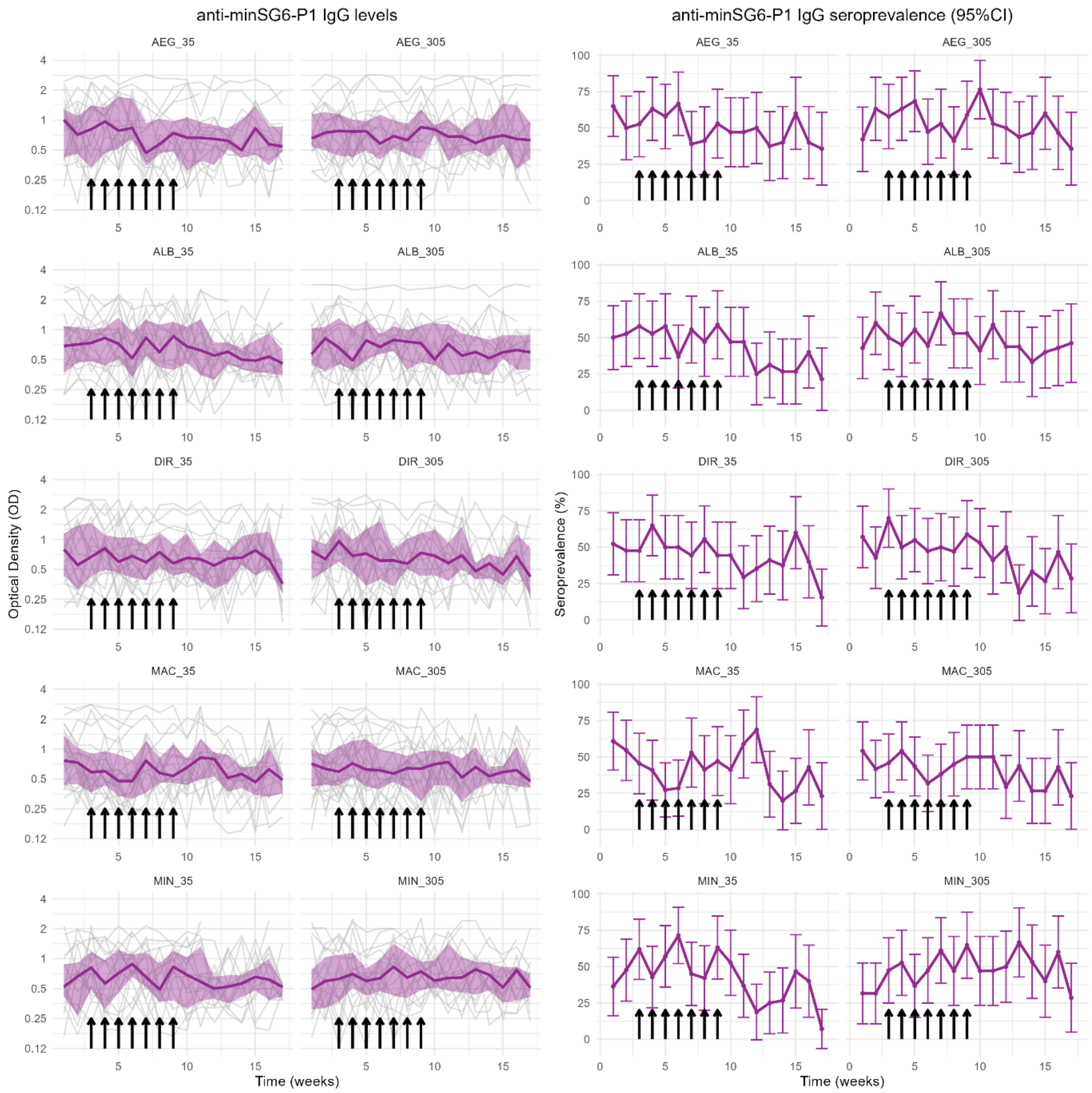

**Figure 3. Observed levels and seroprevalence of antibodies against minSG6-P1 over time, by intervention group.** Left panel shows spaghetti plots of the individual-level anti-minSG6-P1 IgG antibody response over time (week), overlayed with the median and interquartile range of anti-minSG6-P1 antibody levels (OD). Right panel shows the seroprevalence and 95% confidence intervals (CI) each week. Panels represent each intervention group (biting species and dose), and arrows indicate weeks of biting exposure.

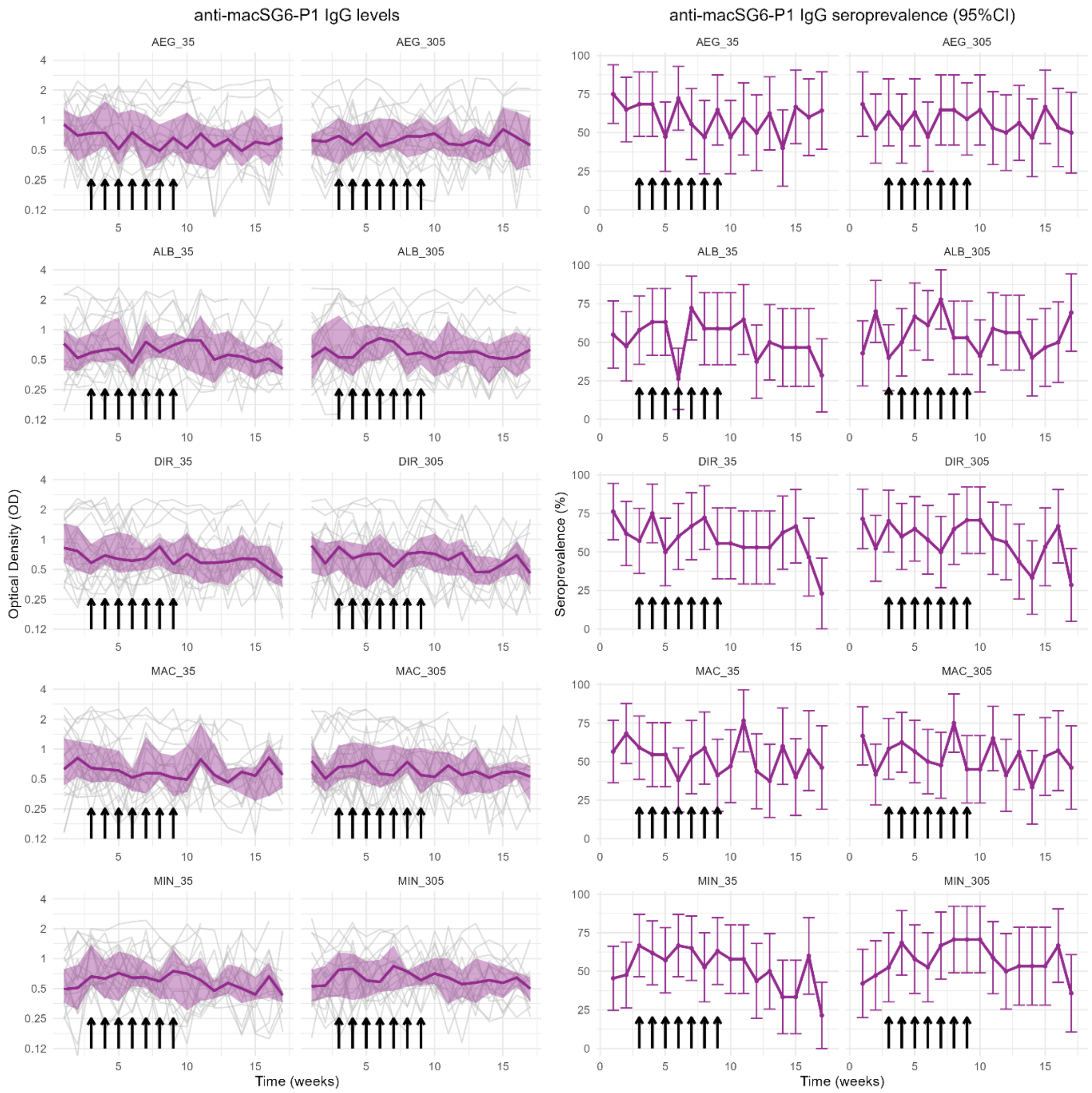

**Figure 4. Observed levels and seroprevalence of antibodies against macSG6-P1 over time, by intervention group.** Left panel shows spaghetti plots of the individual-level anti-macSG6-P1 IgG antibody response over time (week), overlayed with the median and interquartile range of anti-macSG6-P1 antibody levels (OD). Right panel shows the seroprevalence and 95% confidence intervals (CI) each week. Panels represent each intervention group (biting species and dose), and arrows indicate weeks of biting exposure.

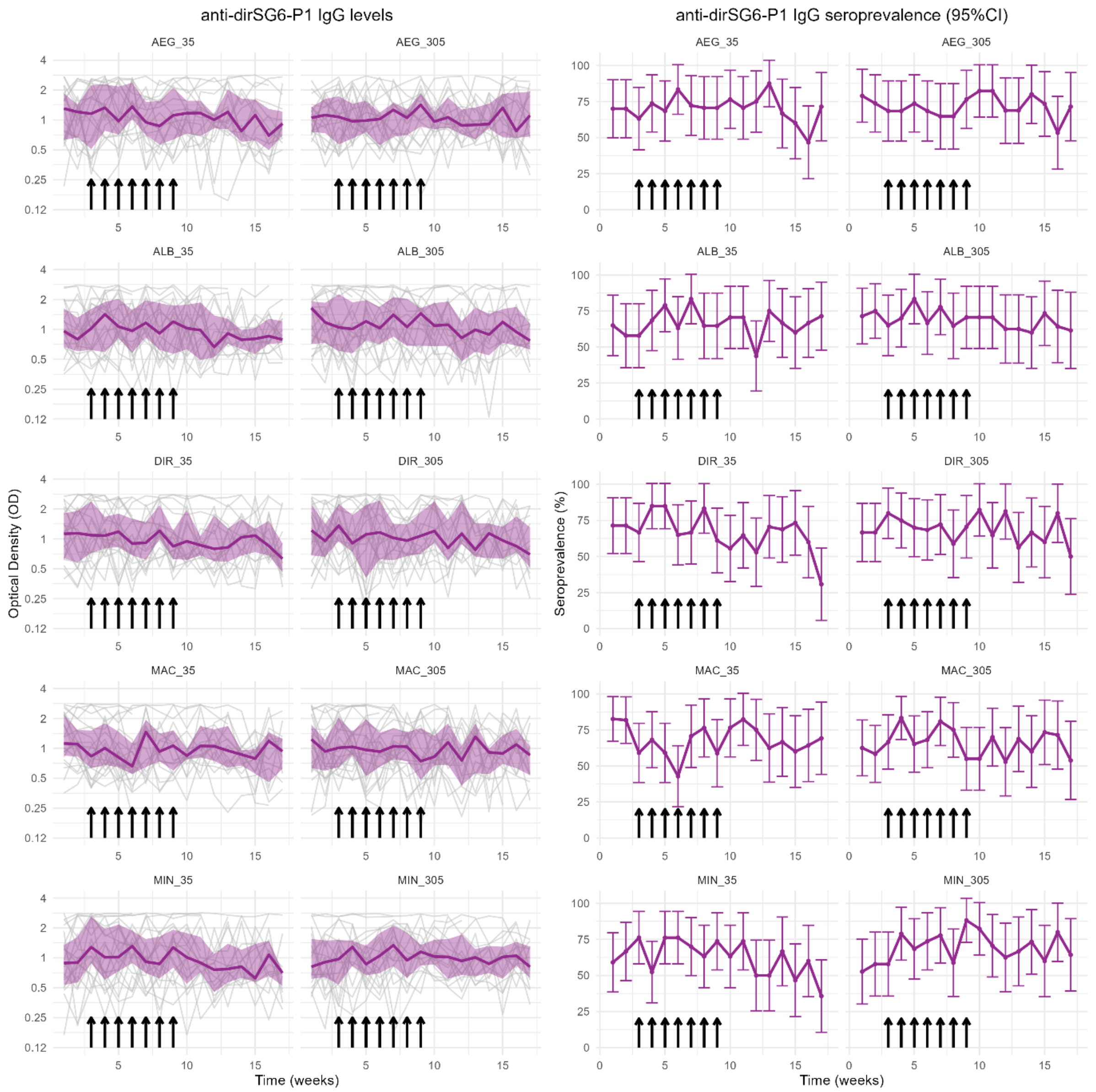

**Figure 5. Observed levels and seroprevalence of antibodies against dirSG6-P1 over time, by intervention group.** Left panel shows spaghetti plots of the individual-level anti-dirSG6-P1 IgG antibody response over time (week), overlayed with the median and interquartile range of anti-dirSG6-P1 antibody levels (OD). Right panel shows the seroprevalence and 95% confidence intervals (CI) each week. Panels represent each intervention group (biting species and dose), and arrows indicate weeks of biting exposure.

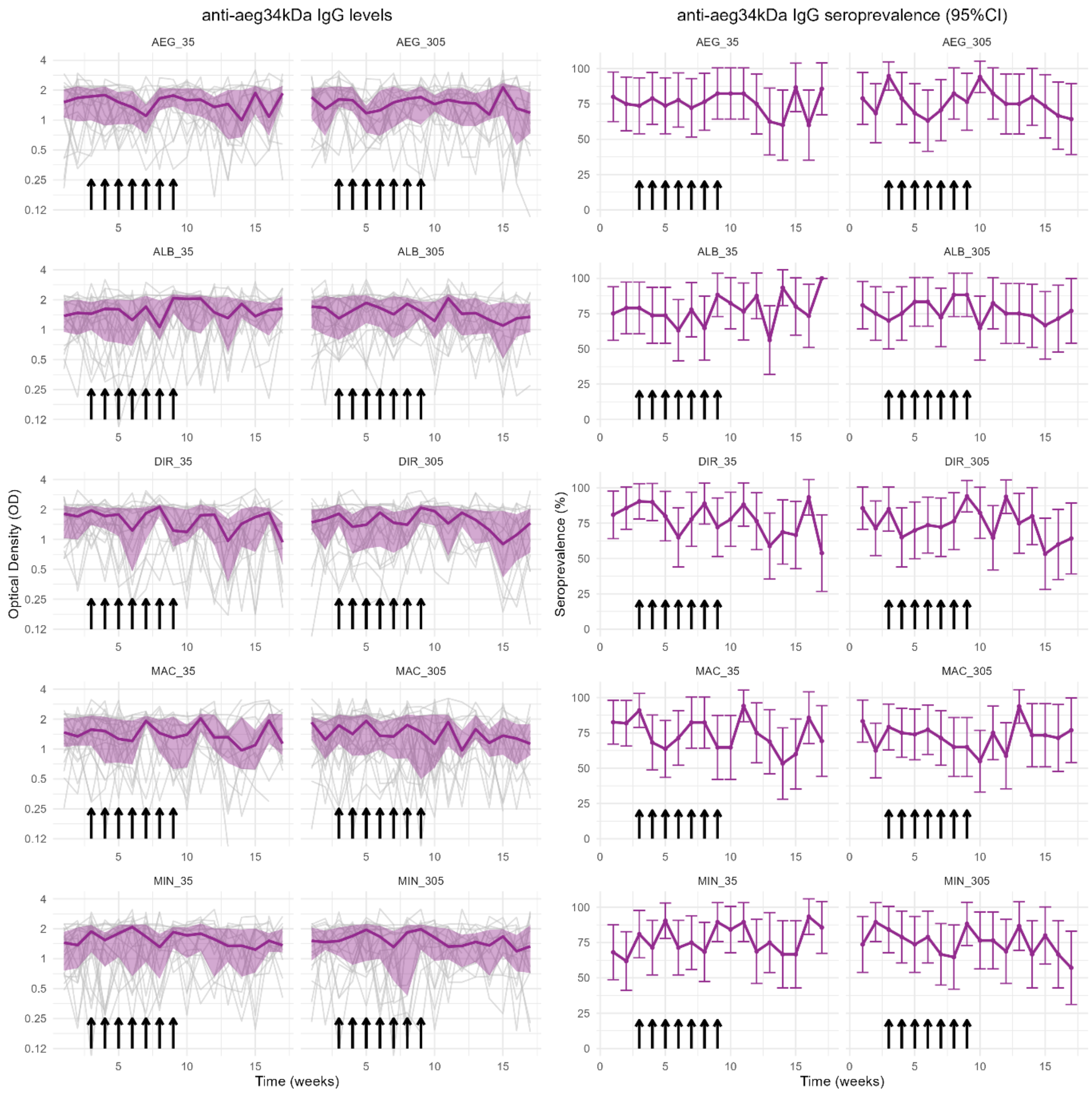

**Figure 6. Observed levels and seroprevalence of antibodies against aeg34kDa over time, by intervention group.** Left panel shows spaghetti plots of the individual-level anti-aeg34kDa IgG antibody response over time (week), overlayed with the median and interquartile range of anti-aeg34kDa antibody levels (OD). Right panel shows the seroprevalence and 95% confidence intervals (CI) each week. Panels represent each intervention group (biting species and dose), and arrows indicate weeks of biting exposure.

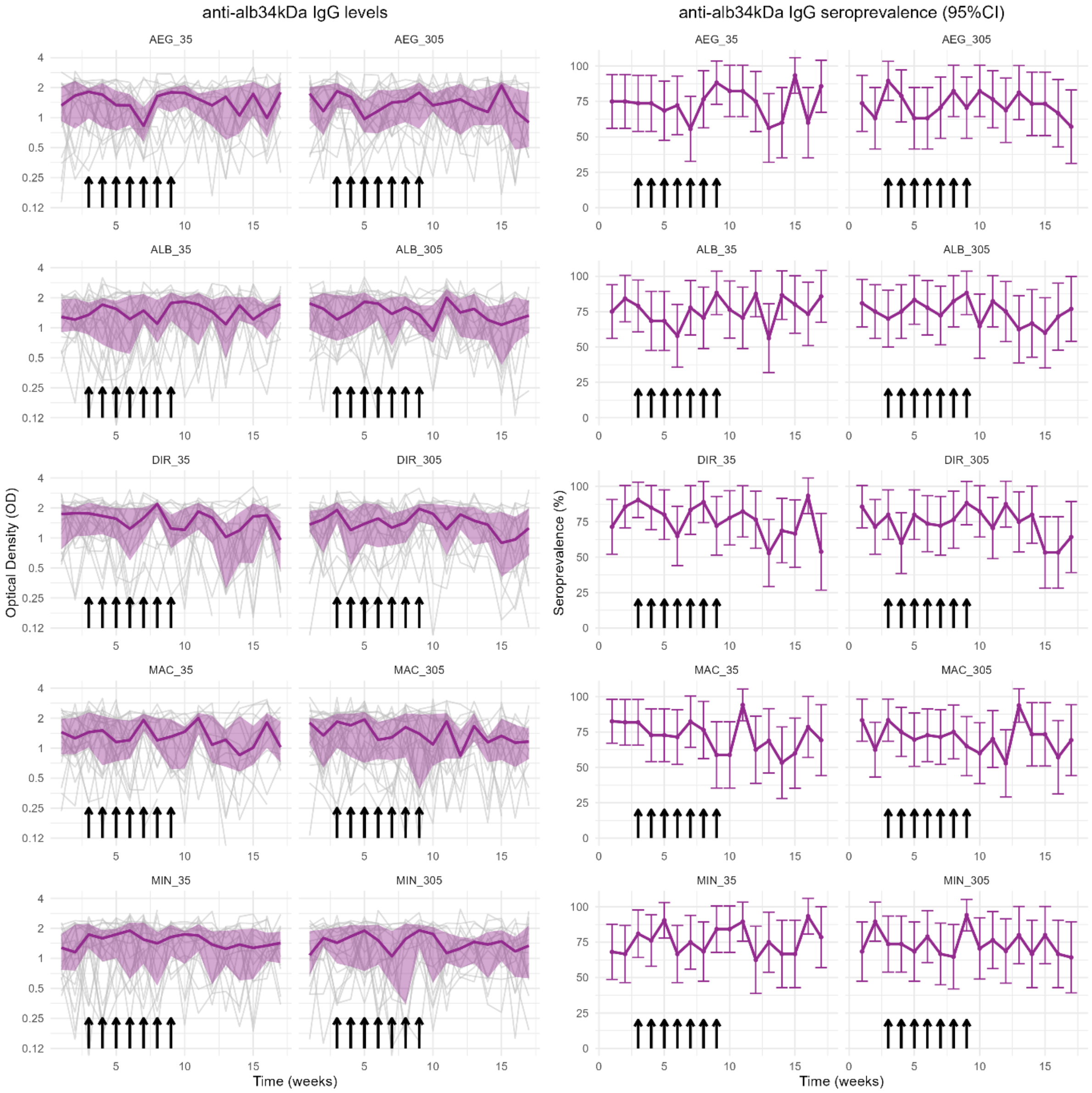

**Figure 7. Observed levels and seroprevalence of antibodies against alb34kDa over time, by intervention group.** Left panel shows spaghetti plots of the individual-level anti-alb34kDa IgG antibody response over time (week), overlayed with the median and interquartile range of anti-alb34kDa antibody levels (OD). Right panel shows the seroprevalence and 95% confidence intervals (CI) each week. Panels represent each intervention group (biting species and dose), and arrows indicate weeks of biting exposure.

### Supplementrary Tables

#### Table 1. Pairwise comparison of SG6-P1 sequences across SG6 orthologs in the Southeast Asian malaria vector species.

| | **gSG6-P1** | **minSG6-P1** | **macSG6-P1** | **dirSG6-P1** |
|---|---|---|---|---|
| **gSG6-P1** | - | 87% (20/23) | 78% (18/23) | 48% (11/23) |
| **minSG6-P1** | 87% (20/23) | - | 78% (18/23) | 57% (13/23) |
| **macSG6-P1** | 78% (18/23) | 78% (18/23) | - | 65% (15/23) |
| **dirSG6-P1** | 48% (11/23) | 57% (13/23) | 65% (15/23) | - |
| | **aeg34kDa** | **alb34kDa** | | |
| **aeg34kDa** | - | 19% (4/21) | | |
| **alb34kDa** | 19% (4/21) | - | | |

#### Table 2. Adverse events

| Intervention group | Adverse Event | Last visit |
| --- | --- | --- |
| *Ae. albopictus*, 305 bites | Became pregnant: withdrawn | 2 |
| *An. dirus*, 35 bites | Atypical skin reaction after the first challenge consisting of hyperpigmentation (self-resolved within 1 month after cession of exposure): withdrawn | 4 |
| *An. dirus*, 305 bites | Experienced pain during venipuncture: self-withdrawn | 9 |
| *An. dirus*, 305 bites | Thrombocytopenia: withdrawn | 3 |
| *An. maculatus*, 35 bites | Palpitation and general weakness (deemed unrelated to the study procedure): withdrawn | 9 |
| *An. maculatus*, 305 bites | Green serum: withdrawn | 18 |
| *An. maculatus*, 305 bites | Grade 1 itching: self-withdrawn | 7 |

**Table 3. Observed values of total IgG antibody responses by intervention group and follow-up period.**

| Group and follow-up period | gSG6-P1 | | dirSG6-P1 | | macSG6-P1 | | minSG6-P1 | | aeg34kDa | | aeg34kDa | |
|---|---|---|---|---|---|---|---|---|---|---|---|---|
| | Level (OD) | Sero-prevalence | Level (OD) | Sero-prevalence | Level (OD) | Sero-prevalence | Level (OD) | Sero-prevalence | Level (OD) | Sero-prevalence | Level (OD) | Sero-prevalence |
| *AEG_35* | | | | | | | | | | | | |
| Baseline | 2.3 (1.54-2.51) | 46/59 (78%) | 1.3 (0.63-1.79) | 40/59 (67.8%) | 0.81 (0.5-1.21) | 41/59 (69.5%) | 0.84 (0.4-1.34) | 33/59 (55.9%) | 1.61 (1.06-2.13) | 45/59 (76.3%) | 1.56 (0.84-2.19) | 44/59 (74.6%) |
| Exposure | 2.24 (1.59-2.5) | 90/108 (83.3%) | 1.13 (0.71-1.84) | 79/108 (73.1%) | 0.67 (0.46-1.24) | 64/108 (59.3%) | 0.74 (0.46-1.26) | 58/108 (53.7%) | 1.48 (0.9-2.05) | 83/108 (76.9%) | 1.41 (0.77-1.95) | 78/108 (72.2%) |
| Post-exposure | 2.01 (1.42-2.44) | 94/125 (75.2%) | 1 (0.65-1.58) | 87/125 (69.6%) | 0.62 (0.36-1) | 70/125 (56%) | 0.65 (0.4-0.96) | 56/125 (44.8%) | 1.58 (0.83-2.06) | 93/125 (74.4%) | 1.55 (0.8-1.98) | 93/125 (74.4%) |
| *AEG_305* | | | | | | | | | | | | |
| Baseline | 2.11 (1.66-2.44) | 46/57 (80.7%) | 1.07 (0.72-1.51) | 42/57 (73.7%) | 0.64 (0.43-0.85) | 35/57 (61.4%) | 0.72 (0.51-1.15) | 31/57 (54.4%) | 1.64 (1.22-2.12) | 46/57 (80.7%) | 1.5 (0.87-2.12) | 43/57 (75.4%) |
| Exposure | 2.08 (1.59-2.43) | 90/108 (83.3%) | 1.05 (0.68-1.52) | 75/108 (69.4%) | 0.64 (0.45-0.99) | 63/108 (58.3%) | 0.75 (0.45-1.13) | 60/108 (55.6%) | 1.47 (0.85-2) | 79/108 (73.1%) | 1.37 (0.76-1.9) | 77/108 (71.3%) |
| Post-exposure | 2.05 (1.55-2.45) | 98/125 (78.4%) | 0.96 (0.71-1.6) | 91/125 (72.8%) | 0.59 (0.4-0.96) | 69/125 (55.2%) | 0.69 (0.44-1.06) | 65/125 (52%) | 1.45 (0.89-2.08) | 96/125 (76.8%) | 1.33 (0.77-2.09) | 91/125 (72.8%) |
| *ALB_35* | | | | | | | | | | | | |
| Baseline | 2.14 (1.5-2.43) | 46/58 (79.3%) | 0.96 (0.63-1.51) | 35/58 (60.3%) | 0.6 (0.4-0.94) | 31/58 (53.4%) | 0.71 (0.44-1.07) | 31/58 (53.4%) | 1.43 (0.9-1.97) | 45/58 (77.6%) | 1.28 (0.88-1.95) | 46/58 (79.3%) |
| Exposure | 2.18 (1.68-2.51) | 94/109 (86.2%) | 1.07 (0.62-1.83) | 77/109 (70.6%) | 0.63 (0.43-1.19) | 62/109 (56.9%) | 0.7 (0.42-1.2) | 56/109 (51.4%) | 1.58 (0.83-2.17) | 80/109 (73.4%) | 1.39 (0.67-2.15) | 78/109 (71.6%) |
| Post-exposure | 1.85 (1.43-2.39) | 96/125 (76.8%) | 0.86 (0.58-1.4) | 82/125 (65.6%) | 0.55 (0.34-0.86) | 60/125 (48%) | 0.55 (0.37-0.91) | 42/125 (33.6%) | 1.6 (1.03-2.14) | 101/125 (80.8%) | 1.52 (0.96-2.02) | 96/125 (76.8%) |
| *ALB_305* | | | | | | | | | | | | |
| Baseline | 2.05 (1.43-2.49) | 46/61 (75.4%) | 1.11 (0.68-1.97) | 43/61 (70.5%) | 0.59 (0.4-1.16) | 31/61 (50.8%) | 0.69 (0.44-1.08) | 31/61 (50.8%) | 1.65 (0.88-2.08) | 46/61 (75.4%) | 1.52 (0.86-2.19) | 46/61 (75.4%) |
| Exposure | 2.14 (1.43-2.43) | 82/108 (75.9%) | 1.17 (0.7-1.97) | 78/108 (72.2%) | 0.69 (0.45-1.04) | 65/108 (60.2%) | 0.73 (0.43-1.08) | 57/108 (52.8%) | 1.59 (0.99-2.09) | 88/108 (81.5%) | 1.52 (0.9-2.04) | 86/108 (79.6%) |
| Post-exposure | 1.86 (1.44-2.46) | 94/123 (76.4%) | 0.99 (0.65-1.66) | 81/123 (65.9%) | 0.57 (0.37-0.88) | 64/123 (52%) | 0.59 (0.39-0.91) | 54/123 (43.9%) | 1.32 (0.85-2.04) | 90/123 (73.2%) | 1.33 (0.73-1.99) | 86/123 (69.9%) |
| *DIR_35* | | | | | | | | | | | | |
| Baseline | 2.14 (1.48-2.47) | 50/63 (79.4%) | 1.12 (0.63-1.97) | 44/63 (69.8%) | 0.67 (0.47-1.43) | 41/63 (65.1%) | 0.67 (0.37-1.34) | 31/63 (49.2%) | 1.88 (1.15-2.18) | 54/63 (85.7%) | 1.76 (1.03-2.18) | 52/63 (82.5%) |
| Exposure | 2.2 (1.52-2.46) | 94/114 (82.5%) | 1.05 (0.72-1.6) | 85/114 (74.6%) | 0.63 (0.47-1.05) | 72/114 (63.2%) | 0.72 (0.45-1.09) | 59/114 (51.8%) | 1.78 (0.99-2.12) | 90/114 (78.9%) | 1.49 (0.91-2.18) | 90/114 (78.9%) |
| Post-exposure | 1.98 (1.42-2.44) | 100/128 (78.1%) | 0.87 (0.61-1.48) | 77/128 (60.2%) | 0.58 (0.38-1) | 67/128 (52.3%) | 0.63 (0.41-1) | 49/128 (38.3%) | 1.43 (0.82-2.05) | 94/128 (73.4%) | 1.35 (0.71-2.01) | 92/128 (71.9%) |
| *DIR_305* | | | | | | | | | | | | |
| Baseline | 2.26 (1.75-2.49) | 52/62 (83.9%) | 1.16 (0.69-2.13) | 44/62 (71%) | 0.78 (0.43-1.07) | 40/62 (64.5%) | 0.75 (0.49-1.19) | 35/62 (56.5%) | 1.7 (1.1-2.05) | 50/62 (80.6%) | 1.68 (1.02-2.02) | 49/62 (79%) |
| Exposure | 2 (1.55-2.44) | 90/111 (81.1%) | 1.06 (0.64-1.75) | 77/111 (69.4%) | 0.67 (0.39-0.93) | 68/111 (61.3%) | 0.69 (0.41-1.08) | 57/111 (51.4%) | 1.51 (0.87-2.14) | 83/111 (74.8%) | 1.47 (0.81-2.13) | 83/111 (74.8%) |
| Post-exposure | 1.92 (1.45-2.44) | 96/125 (76.8%) | 0.96 (0.64-1.72) | 85/125 (68%) | 0.59 (0.37-0.85) | 65/125 (52%) | 0.56 (0.39-0.95) | 47/125 (37.6%) | 1.55 (0.83-2) | 90/125 (72%) | 1.36 (0.69-2.01) | 89/125 (71.2%) |
| *MAC_35* | | | | | | | | | | | | |
| Baseline | 1.99 (1.58-2.51) | 55/67 (82.1%) | 1.05 (0.72-1.73) | 50/67 (74.6%) | 0.67 (0.47-1.22) | 41/67 (61.2%) | 0.71 (0.4-1.08) | 36/67 (53.7%) | 1.47 (1.05-2.1) | 57/67 (85.1%) | 1.35 (0.89-2.06) | 55/67 (82.1%) |
| Exposure | 1.92 (1.37-2.44) | 86/116 (74.1%) | 0.92 (0.6-1.61) | 72/116 (62.1%) | 0.56 (0.41-0.99) | 58/116 (50%) | 0.57 (0.37-0.97) | 45/116 (38.8%) | 1.54 (0.77-2.06) | 83/116 (71.6%) | 1.46 (0.7-2) | 84/116 (72.4%) |
| Post-exposure | 1.95 (1.48-2.41) | 96/123 (78%) | 0.91 (0.65-1.28) | 86/123 (69.9%) | 0.58 (0.37-0.92) | 63/123 (51.2%) | 0.59 (0.38-0.84) | 49/123 (39.8%) | 1.4 (0.81-2.1) | 88/123 (71.5%) | 1.36 (0.72-2.05) | 84/123 (68.3%) |

| | | | | | | | | | | | | |
|---|---|---|---|---|---|---|---|---|---|---|---|---|
| *MAC_305* | | | | | | | | | | | | |
| Baseline | 2.14 (1.39-2.48) | 53/72 (73.6%) | 0.98 (0.64-1.66) | 45/72 (62.5%) | 0.57 (0.45-1.07) | 40/72 (55.6%) | 0.65 (0.42-0.97) | 34/72 (47.2%) | 1.73 (0.96-2.12) | 54/72 (75%) | 1.65 (0.91-2.02) | 55/72 (76.4%) |
| Exposure | 1.9 (1.39-2.48) | 97/130 (74.6%) | 0.98 (0.67-1.51) | 93/130 (71.5%) | 0.64 (0.42-1.13) | 73/130 (56.2%) | 0.64 (0.41-1.1) | 57/130 (43.8%) | 1.46 (0.82-2.15) | 93/130 (71.5%) | 1.43 (0.71-2.21) | 93/130 (71.5%) |
| Post-exposure | 2.01 (1.46-2.32) | 101/130 (77.7%) | 0.91 (0.55-1.42) | 82/130 (63.1%) | 0.56 (0.39-0.87) | 65/130 (50%) | 0.57 (0.4-0.93) | 49/130 (37.7%) | 1.31 (0.81-1.96) | 93/130 (71.5%) | 1.26 (0.76-1.94) | 89/130 (68.5%) |
| *MIN_35* | | | | | | | | | | | | |
| Baseline | 1.96 (1.59-2.5) | 51/64 (79.7%) | 0.99 (0.61-1.77) | 43/64 (67.2%) | 0.57 (0.4-1.01) | 34/64 (53.1%) | 0.64 (0.4-1.02) | 31/64 (48.4%) | 1.55 (0.82-2.07) | 45/64 (70.3%) | 1.38 (0.77-2.1) | 46/64 (71.9%) |
| Exposure | 2.11 (1.7-2.5) | 99/121 (81.8%) | 1.1 (0.68-1.88) | 83/121 (68.6%) | 0.66 (0.43-1.07) | 74/121 (61.2%) | 0.71 (0.39-1.11) | 65/121 (53.7%) | 1.64 (0.95-2.15) | 94/121 (77.7%) | 1.56 (0.89-2.2) | 93/121 (76.9%) |
| Post-exposure | 1.81 (1.43-2.33) | 99/129 (76.7%) | 0.82 (0.57-1.37) | 73/129 (56.6%) | 0.51 (0.37-0.81) | 59/129 (45.7%) | 0.54 (0.39-0.8) | 42/129 (32.6%) | 1.51 (0.97-2.08) | 102/129 (79.1%) | 1.41 (0.86-1.96) | 100/129 (77.5%) |
| *MIN_305* | | | | | | | | | | | | |
| Baseline | 1.83 (1.29-2.46) | 38/57 (66.7%) | 0.91 (0.56-1.54) | 32/57 (56.1%) | 0.53 (0.38-0.86) | 27/57 (47.4%) | 0.55 (0.33-1.03) | 21/57 (36.8%) | 1.51 (1.12-2.03) | 47/57 (82.5%) | 1.45 (0.95-1.97) | 44/57 (77.2%) |
| Exposure | 2.16 (1.71-2.46) | 88/109 (80.7%) | 1.1 (0.71-1.68) | 81/109 (74.3%) | 0.72 (0.48-1) | 70/109 (64.2%) | 0.69 (0.42-1.03) | 56/109 (51.4%) | 1.77 (0.86-2.17) | 82/109 (75.2%) | 1.66 (0.8-2.15) | 81/109 (74.3%) |
| Post-exposure | 1.95 (1.61-2.4) | 100/124 (80.6%) | 1.01 (0.68-1.37) | 87/124 (70.2%) | 0.61 (0.42-0.86) | 69/124 (55.6%) | 0.66 (0.43-0.92) | 61/124 (49.2%) | 1.37 (0.82-2) | 90/124 (72.6%) | 1.31 (0.77-1.84) | 89/124 (71.8%) |

Data are median (IQR) for anti-salivary antibody levels (Optical Density ($OD_{405nm}$)), or n/N (%) for anti-salivary antibody seroprevalence. Antibody levels were calculated using the average OD value of the 3 baseline samples collated by participant.

**Table 4. Effect of mosquito biting exposure period (across all intervention groups) on anti-salivary antibody levels.**

| | gSG6-P1 | | | minSG6-P1 | | | macSG6-P1 | | | dirSG6-P1 | | | aeg34kDa | | | alb34kDa | | |
|---|---|---|---|---|---|---|---|---|---|---|---|---|---|---|---|---|---|---|
| **Variable** | **GMR** | **95%CI** | ***p*** | **GMR** | **95%CI** | ***p*** | **GMR** | **95%CI** | ***p*** | **GMR** | **95%CI** | ***p*** | **GMR** | **95%CI** | ***p*** | **GMR** | **95%CI** | ***p*** |
| Time (Days) | 0.999 | 0.998 - 0.999 | 0.006 | 0.998 | 0.997 - 0.999 | 0.005 | 0.998 | 0.997 - 0.999 | 0.002 | 0.998 | 0.996 - 0.999 | <0.001 | 0.999 | 0.997 - 1.000 | 0.061 | 0.998 | 0.997 - 0.999 | 0.031 |
| Intervention Period | | | | | | | | | | | | | | | | | | |
| *Baseline* | Ref. | | | Ref. | | | Ref. | | | Ref. | | | Ref. | | | Ref. | | |
| *Exposure* | 1.045 | 1.007 - 1.084 | 0.020 | 1.040 | 0.977 - 1.107 | 0.216 | 1.046 | 0.985 - 1.111 | 0.144 | 1.061 | 0.999 - 1.127 | 0.053 | 1.006 | 0.935 - 1.083 | 0.863 | 1.014 | 0.941 - 1.091 | 0.721 |
| *Post-Exposure* | 1.060 | 0.989 - 1.136 | 0.099 | 1.044 | 0.933 - 1.167 | 0.452 | 1.046 | 0.941 - 1.161 | 0.405 | 1.100 | 0.986 - 1.228 | 0.089 | 1.042 | 0.927 - 1.171 | 0.494 | 1.057 | 0.937 - 1.193 | 0.368 |

**Note.** Data are given as geometric mean ratio (GMR), 95% confidence interval (95%CI), p-value (*p*), estimated from generalised estimating equations (GEEs) analysis (n=206 participants, of note 4 participants that provided <2 antibody measurements were excluded from analysis due to the specification of an autoregressive correlation structure) of the effect of mosquito biting intervention and time (days) on the $\log_2$(OD) levels of IgG antibodies against species-specific *Anopheles* and *Aedes* salivary antigens (adjusted by age (years) and sex).

**Table 5. Time-dependent effect of mosquito biting exposure period (across all intervention groups) on boosting and decay of anti-salivary antibodies.**

| | gSG6-P1 | | | minSG6-P1 | | | macSG6-P1 | | | dirSG6-P1 | | | aeg34kDa | | | alb34kDa | | |
|---|---|---|---|---|---|---|---|---|---|---|---|---|---|---|---|---|---|---|
| **Variable** | **GMR** | **95%CI** | ***p*** | **GMR** | **95%CI** | ***p*** | **GMR** | **95%CI** | ***p*** | **GMR** | **95%CI** | ***p*** | **GMR** | **95%CI** | ***p*** | **GMR** | **95%CI** | ***p*** |
| Time (Days) | 1.002 | 0.998 - 1.006 | 0.381 | 1.003 | 0.997 - 1.009 | 0.337 | 1.003 | 0.997 - 1.009 | 0.266 | 1.000 | 0.994 - 1.006 | 0.949 | 1.003 | 0.996 - 1.010 | 0.427 | 1.004 | 0.997 - 1.011 | 0.298 |
| Time X Intervention Period | | | | | | | | | | | | | | | | | | |
| *Baseline* | Ref. | | | Ref. | | | Ref. | | | Ref. | | | Ref. | | | Ref. | | |
| *Exposure* | 0.999 | 0.998 - 1.001 | 0.333 | 1.000 | 0.997 - 1.002 | 0.782 | 0.999 | 0.997 - 1.002 | 0.623 | 0.999 | 0.997 - 1.002 | 0.457 | 1.001 | 0.998 - 1.004 | 0.572 | 1.000 | 0.997 - 1.003 | 0.799 |
| *Post-Exposure* | 0.998 | 0.997 - 0.999 | 0.002 | 0.997 | 0.995 - 0.998 | <0.001 | 0.996 | 0.995 - 0.998 | <0.001 | 0.997 | 0.995 - 0.998 | <0.001 | 0.997 | 0.996 - 0.999 | 0.002 | 0.997 | 0.995 - 0.999 | 0.001 |

**Note.** Data are given as geometric mean ratio (GMR), 95% confidence interval (95%CI), p-value (*p*), estimated from generalised estimating equations (GEEs) analysis (n=206 participants, of note 4 participants that provided <2 antibody measurements were excluded from analysis due to the specification of an autoregressive correlation structure) of the effect of mosquito biting intervention and time (days) on the $\log_2$(OD) levels of IgG antibodies against species-specific *Anopheles* and *Aedes* salivary antigens (adjusted by age (years) and sex), and includes an interaction term between time and the mosquito biting intervention period to allow changes in the antibody levels over time to be dependent on the intervention.

**Table 6. Effect of mosquito biting exposure period, modified by intervention group (species and dose), on anti-salivary antibody levels.**

| | **gSG6-P1** | | | **minSG6-P1** | | | **macSG6-P1** | | | **dirSG6-P1** | | | **aeg34kDa** | | | **alb34kDa** | | |
|---|---|---|---|---|---|---|---|---|---|---|---|---|---|---|---|---|---|---|
| **Variable** | **GMR** | **95%CI** | ***p*** | **GMR** | **95%CI** | ***p*** | **GMR** | **95%CI** | ***p*** | **GMR** | **95%CI** | ***p*** | **GMR** | **95%CI** | ***p*** | **GMR** | **95%CI** | ***p*** |
| Time (Days) | 0.999 | 0.998 - 0.999 | 0.005 | 0.998 | 0.997 - 0.999 | 0.005 | 0.998 | 0.997 - 0.999 | 0.002 | 0.998 | 0.996 - 0.999 | <0.001 | 0.999 | 0.997 - 1.000 | 0.048 | 0.998 | 0.997 - 0.999 | 0.025 |
| Exposure period X Intervention Group | | | | | | | | | | | | | | | | | | |
| *Baseline* | Ref. | | | Ref. | | | Ref. | | | Ref. | | | Ref. | | | Ref. | | |
| *AEG_35* | 1.058 | 0.984 - 1.137 | 0.130 | 1.110 | 0.972 - 1.268 | 0.125 | 1.077 | 0.940 - 1.234 | 0.287 | 1.116 | 0.986 - 1.263 | 0.083 | 1.053 | 0.868 - 1.277 | 0.601 | 1.030 | 0.849 - 1.250 | 0.766 |
| *AEG_305* | 1.020 | 0.923 - 1.127 | 0.699 | 1.007 | 0.853 - 1.187 | 0.938 | 1.078 | 0.902 - 1.288 | 0.409 | 1.063 | 0.893 - 1.267 | 0.491 | 0.935 | 0.779 - 1.121 | 0.468 | 0.961 | 0.808 - 1.142 | 0.649 |
| *ALB_35* | 1.077 | 0.982 - 1.182 | 0.116 | 1.018 | 0.876 - 1.183 | 0.817 | 1.093 | 0.957 - 1.249 | 0.189 | 1.146 | 0.995 - 1.321 | 0.059 | 1.033 | 0.852 - 1.253 | 0.741 | 1.041 | 0.859 - 1.262 | 0.683 |
| *ALB_305* | 1.038 | 0.955 - 1.129 | 0.380 | 1.015 | 0.878 - 1.173 | 0.842 | 1.063 | 0.908 - 1.243 | 0.447 | 1.033 | 0.883 - 1.208 | 0.687 | 1.130 | 0.971 - 1.315 | 0.115 | 1.114 | 0.958 - 1.295 | 0.161 |
| *DIR_35* | 1.061 | 0.974 - 1.157 | 0.176 | 1.029 | 0.865 - 1.224 | 0.748 | 1.016 | 0.882 - 1.171 | 0.826 | 1.036 | 0.890 - 1.207 | 0.647 | 0.913 | 0.804 - 1.036 | 0.159 | 0.918 | 0.805 - 1.046 | 0.199 |
| *DIR_305* | 0.973 | 0.891 - 1.063 | 0.542 | 0.946 | 0.819 - 1.092 | 0.445 | 0.989 | 0.863 - 1.133 | 0.873 | 0.917 | 0.799 - 1.051 | 0.214 | 0.997 | 0.810 - 1.227 | 0.979 | 0.991 | 0.801 - 1.228 | 0.938 |
| *MAC_35* | 0.983 | 0.901 - 1.073 | 0.708 | 0.998 | 0.885 - 1.125 | 0.973 | 0.955 | 0.834 - 1.093 | 0.502 | 1.024 | 0.891 - 1.177 | 0.738 | 0.954 | 0.809 - 1.124 | 0.570 | 0.987 | 0.829 - 1.176 | 0.885 |
| *MAC_305* | 1.024 | 0.941 - 1.115 | 0.583 | 1.081 | 0.950 - 1.229 | 0.236 | 1.025 | 0.912 - 1.153 | 0.675 | 1.057 | 0.933 - 1.197 | 0.386 | 0.990 | 0.810 - 1.211 | 0.923 | 0.973 | 0.793 - 1.193 | 0.794 |
| *MIN_35* | 1.104 | 0.978 - 1.247 | 0.110 | 0.989 | 0.846 - 1.157 | 0.892 | 1.028 | 0.880 - 1.199 | 0.730 | 1.073 | 0.906 - 1.269 | 0.414 | 1.088 | 0.892 - 1.326 | 0.407 | 1.120 | 0.937 - 1.339 | 0.214 |
| *MIN_305* | 1.138 | 1.028 - 1.260 | 0.012 | 1.256 | 1.019 - 1.548 | 0.033 | 1.195 | 0.996 - 1.434 | 0.055 | 1.198 | 0.999 - 1.437 | 0.052 | 1.034 | 0.863 - 1.238 | 0.719 | 1.060 | 0.873 - 1.287 | 0.559 |
| Post-exposure period X Intervention Group | | | | | | | | | | | | | | | | | | |
| *Baseline* | Ref. | | | Ref. | | | Ref. | | | Ref. | | | Ref. | | | Ref. | | |
| *AEG_35* | 1.026 | 0.905 - 1.162 | 0.693 | 1.062 | 0.873 - 1.293 | 0.546 | 1.061 | 0.871 - 1.292 | 0.557 | 1.158 | 0.974 - 1.377 | 0.098 | 1.124 | 0.916 - 1.380 | 0.262 | 1.170 | 0.942 - 1.454 | 0.156 |
| *AEG_305* | 1.052 | 0.951 - 1.165 | 0.325 | 1.100 | 0.912 - 1.327 | 0.318 | 1.134 | 0.937 - 1.372 | 0.195 | 1.141 | 0.925 - 1.407 | 0.219 | 1.006 | 0.806 - 1.255 | 0.959 | 1.013 | 0.809 - 1.270 | 0.908 |
| *ALB_35* | 1.044 | 0.939 - 1.160 | 0.426 | 0.953 | 0.797 - 1.14 | 0.597 | 1.041 | 0.886 - 1.222 | 0.626 | 1.147 | 0.969 - 1.357 | 0.111 | 1.223 | 1.037 - 1.442 | 0.017 | 1.227 | 1.034 - 1.456 | 0.019 |
| *ALB_305* | 1.047 | 0.936 - 1.170 | 0.422 | 1.010 | 0.843 - 1.209 | 0.917 | 1.071 | 0.896 - 1.280 | 0.452 | 1.055 | 0.876 - 1.27 | 0.573 | 1.045 | 0.859 - 1.270 | 0.660 | 1.058 | 0.862 - 1.298 | 0.590 |
| *DIR_35* | 1.080 | 0.961 - 1.214 | 0.196 | 1.049 | 0.846 - 1.302 | 0.662 | 1.013 | 0.840 - 1.222 | 0.892 | 1.075 | 0.883 - 1.308 | 0.473 | 0.882 | 0.724 - 1.074 | 0.213 | 0.904 | 0.734 - 1.113 | 0.341 |
| *DIR_305* | 0.994 | 0.889 - 1.111 | 0.915 | 0.950 | 0.777 - 1.161 | 0.614 | 0.983 | 0.820 - 1.178 | 0.850 | 1.054 | 0.870 - 1.278 | 0.590 | 0.996 | 0.763 - 1.301 | 0.979 | 0.986 | 0.747 - 1.302 | 0.921 |
| *MAC_35* | 1.069 | 0.953 - 1.198 | 0.256 | 0.980 | 0.812 - 1.182 | 0.829 | 0.983 | 0.826 - 1.170 | 0.849 | 1.042 | 0.864 - 1.255 | 0.669 | 0.999 | 0.831 - 1.201 | 0.992 | 1.027 | 0.831 - 1.270 | 0.802 |
| *MAC_305* | 1.082 | 0.928 - 1.262 | 0.316 | 1.093 | 0.876 - 1.364 | 0.431 | 0.982 | 0.803 - 1.201 | 0.859 | 1.087 | 0.892 - 1.323 | 0.408 | 1.011 | 0.817 - 1.252 | 0.918 | 1.004 | 0.806 - 1.251 | 0.970 |
| *MIN_35* | 1.056 | 0.917 - 1.217 | 0.446 | 1.004 | 0.817 - 1.236 | 0.966 | 1.017 | 0.847 - 1.221 | 0.857 | 1.026 | 0.827 - 1.273 | 0.814 | 1.184 | 0.951 - 1.474 | 0.131 | 1.187 | 0.962 - 1.464 | 0.110 |
| *MIN_305* | 1.182 | 1.051 - 1.330 | 0.005 | 1.290 | 1.025 - 1.624 | 0.030 | 1.251 | 1.027 - 1.524 | 0.026 | 1.272 | 1.032 - 1.569 | 0.024 | 1.072 | 0.889 - 1.292 | 0.466 | 1.119 | 0.915 - 1.370 | 0.274 |

**Note.** Data are given as geometric mean ratio (GMR), 95% confidence interval (95%CI), p-value (*p*), estimated from generalised estimating equations (GEEs) analysis (n=206 participants, of note 4 participants that provided <2 antibody measurements were excluded from analysis due to the specification of an autoregressive correlation structure) of the effect of mosquito biting intervention and time (days) on the $log_2$(OD) levels of IgG antibodies against species-specific *Anopheles* and *Aedes* salivary antigens (adjusted by age (years) and sex), and includes an interaction term between the intervention group (i.e. biting species and dose) and the mosquito biting intervention period.

**Table 7. Effect of mosquito biting exposure, modified by intervention group (species and dose), on anti-salivary antibody seroprevalence.**

| | gSG6-P1 | | | minSG6-P1 | | | macSG6-P1 | | | dirSG6-P1 | | | aeg34kDa | | | alb34kDa | | |
|---|---|---|---|---|---|---|---|---|---|---|---|---|---|---|---|---|---|---|
| **Variable** | **OR** | **95%CI** | ***p*** | **OR** | **95%CI** | ***p*** | **OR** | **95%CI** | ***p*** | **OR** | **95%CI** | ***p*** | **OR** | **95%CI** | ***p*** | **OR** | **95%CI** | ***p*** |
| Time (Days) | 0.988 | 0.982 - 0.994 | <0.001 | 0.991 | 0.986 - 0.996 | 0.001 | 0.993 | 0.988 - 0.998 | 0.008 | 0.990 | 0.984 - 0.996 | <0.001 | 0.997 | 0.992 - 1.002 | 0.279 | 0.998 | 0.992 - 1.003 | 0.350 |
| Exposure period X Intervention Group | | | | | | | | | | | | | | | | | | |
| *Baseline* | Ref. | | | Ref. | | | Ref. | | | Ref. | | | Ref. | | | Ref. | | |
| *AEG_35* | 2.278 | 1.272 - 4.081 | 0.006 | 1.271 | 0.884 - 1.829 | 0.195 | 0.850 | 0.506 - 1.429 | 0.54 | 1.787 | 0.989 - 3.231 | 0.055 | 1.151 | 0.518 - 2.561 | 0.730 | 0.974 | 0.458 - 2.069 | 0.945 |
| *AEG_305* | 1.802 | 0.662 - 4.903 | 0.249 | 1.389 | 0.708 - 2.726 | 0.339 | 1.009 | 0.527 - 1.933 | 0.977 | 1.107 | 0.504 - 2.429 | 0.800 | 0.687 | 0.250 - 1.887 | 0.466 | 0.855 | 0.424 - 1.725 | 0.662 |
| *ALB_35* | 2.360 | 0.985 - 5.651 | 0.054 | 1.140 | 0.668 - 1.944 | 0.631 | 1.369 | 0.756 - 2.479 | 0.300 | 2.053 | 1.085 - 3.885 | 0.027 | 0.911 | 0.430 - 1.930 | 0.807 | 0.735 | 0.329 - 1.646 | 0.455 |
| *ALB_305* | 1.460 | 0.690 - 3.090 | 0.322 | 1.445 | 0.751 - 2.780 | 0.270 | 1.939 | 1.099 - 3.419 | 0.022 | 1.559 | 0.750 - 3.241 | 0.234 | 1.614 | 0.684 - 3.805 | 0.274 | 1.412 | 0.693 - 2.880 | 0.342 |
| *DIR_35* | 1.779 | 0.870 - 3.637 | 0.115 | 1.581 | 0.781 - 3.199 | 0.203 | 1.223 | 0.632 - 2.367 | 0.551 | 1.846 | 0.959 - 3.553 | 0.067 | 0.705 | 0.364 - 1.367 | 0.301 | 0.865 | 0.478 - 1.565 | 0.632 |
| *DIR_305* | 1.187 | 0.571 - 2.467 | 0.647 | 0.952 | 0.535 - 1.691 | 0.866 | 1.021 | 0.606 - 1.721 | 0.938 | 1.264 | 0.720 - 2.218 | 0.415 | 0.752 | 0.331 - 1.707 | 0.495 | 0.815 | 0.361 - 1.841 | 0.622 |
| *MAC_35* | 0.962 | 0.454 - 2.041 | 0.920 | 0.772 | 0.424 - 1.406 | 0.397 | 0.781 | 0.404 - 1.511 | 0.462 | 0.817 | 0.395 - 1.690 | 0.585 | 0.484 | 0.220 - 1.062 | 0.070 | 0.630 | 0.299 - 1.326 | 0.224 |
| *MAC_305* | 1.567 | 0.925 - 2.656 | 0.095 | 1.203 | 0.705 - 2.050 | 0.498 | 1.252 | 0.857 - 1.830 | 0.245 | 2.131 | 1.300 - 3.494 | 0.003 | 0.923 | 0.493 - 1.730 | 0.803 | 0.831 | 0.419 - 1.651 | 0.597 |
| *MIN_35* | 1.647 | 0.742 - 3.655 | 0.220 | 1.422 | 0.878 - 2.303 | 0.153 | 1.575 | 0.828 - 2.997 | 0.166 | 1.390 | 0.602 - 3.211 | 0.441 | 1.612 | 0.813 - 3.197 | 0.172 | 1.421 | 0.766 - 2.636 | 0.265 |
| *MIN_305* | 2.938 | 1.709 - 5.050 | <0.001 | 2.325 | 1.238 - 4.367 | 0.009 | 2.455 | 1.397 - 4.314 | 0.002 | 3.177 | 1.672 - 6.033 | <0.001 | 0.731 | 0.326 - 1.639 | 0.447 | 0.953 | 0.527 - 1.726 | 0.875 |
| Post-exposure period X Intervention Group | | | | | | | | | | | | | | | | | | |
| *Baseline* | Ref. | | | Ref. | | | Ref. | | | Ref. | | | Ref. | | | Ref. | | |
| *AEG_35* | 2.410 | 0.998 - 5.820 | 0.051 | 1.304 | 0.735 - 2.313 | 0.365 | 0.985 | 0.468 - 2.074 | 0.969 | 2.532 | 1.313 - 4.883 | 0.006 | 1.175 | 0.526 - 2.625 | 0.695 | 1.222 | 0.542 - 2.754 | 0.630 |
| *AEG_305* | 2.347 | 0.978 - 5.635 | 0.056 | 1.968 | 0.940 - 4.120 | 0.073 | 1.300 | 0.611 - 2.766 | 0.496 | 2.171 | 0.963 - 4.895 | 0.062 | 0.985 | 0.335 - 2.898 | 0.978 | 1.039 | 0.385 - 2.806 | 0.939 |
| *ALB_35* | 2.322 | 0.964 - 5.593 | 0.060 | 0.856 | 0.440 - 1.667 | 0.648 | 1.359 | 0.650 - 2.841 | 0.415 | 2.809 | 1.225 - 6.439 | 0.015 | 1.628 | 0.829 - 3.197 | 0.157 | 1.089 | 0.525 - 2.257 | 0.819 |
| *ALB_305* | 2.899 | 1.141 - 7.364 | 0.025 | 1.581 | 0.735 - 3.401 | 0.241 | 1.971 | 1.032 - 3.767 | 0.040 | 1.937 | 0.818 - 4.586 | 0.133 | 1.148 | 0.529 - 2.493 | 0.727 | 0.940 | 0.436 - 2.024 | 0.873 |
| *DIR_35* | 2.538 | 1.024 - 6.290 | 0.044 | 1.378 | 0.640 - 2.967 | 0.413 | 1.065 | 0.500 - 2.268 | 0.870 | 1.541 | 0.705 - 3.366 | 0.278 | 0.606 | 0.242 - 1.521 | 0.286 | 0.674 | 0.303 - 1.497 | 0.332 |
| *DIR_305* | 1.679 | 0.676 - 4.166 | 0.264 | 0.861 | 0.419 - 1.770 | 0.684 | 0.992 | 0.519 - 1.895 | 0.981 | 1.999 | 0.939 - 4.254 | 0.072 | 0.755 | 0.298 - 1.913 | 0.554 | 0.772 | 0.315 - 1.894 | 0.572 |
| *MAC_35* | 2.124 | 0.791 - 5.705 | 0.135 | 1.127 | 0.583 - 2.180 | 0.722 | 1.138 | 0.567 - 2.282 | 0.717 | 1.893 | 0.824 - 4.348 | 0.133 | 0.560 | 0.254 - 1.233 | 0.150 | 0.582 | 0.243 - 1.392 | 0.224 |
| *MAC_305* | 3.422 | 1.486 - 7.881 | 0.004 | 1.400 | 0.691 - 2.837 | 0.350 | 1.382 | 0.704 - 2.714 | 0.348 | 2.391 | 1.109 - 5.154 | 0.026 | 1.055 | 0.488 - 2.282 | 0.891 | 0.806 | 0.377 - 1.724 | 0.579 |
| *MIN_35* | 2.146 | 0.899 - 5.122 | 0.085 | 0.990 | 0.480 - 2.043 | 0.979 | 1.225 | 0.569 - 2.636 | 0.603 | 1.407 | 0.537 - 3.685 | 0.487 | 2.044 | 0.920 - 4.542 | 0.079 | 1.678 | 0.764 - 3.684 | 0.197 |
| *MIN_305* | 5.539 | 2.728 - 11.245 | <0.001 | 3.074 | 1.506 - 6.273 | 0.002 | 2.459 | 1.241 - 4.875 | 0.010 | 4.280 | 2.222 - 8.245 | <0.001 | 0.735 | 0.321 - 1.682 | 0.466 | 0.928 | 0.467 - 1.845 | 0.831 |

**Note.** Data are given as estimated odds ratio (OR), 95% confidence interval (95%CI), p-value (*p*), estimated from generalised estimating equations (GEEs) analysis (n=206 participants, of note 4 participants that provided <2 antibody measurements were excluded from analysis due to the specification of an autoregressive correlation structure) of the effect of mosquito biting intervention and time (days) on the seropositivity of IgG antibodies against species-specific *Anopheles* and *Aedes* salivary antigens (adjusted by age (years) and sex), and includes an interaction term between the intervention group (i.e. biting species and dose) and the mosquito biting intervention period.
